# An Integrated Germline and Somatic Genomic Model Improves Risk Prediction for Coronary Artery Disease

**DOI:** 10.1101/2024.11.01.24316612

**Authors:** Xiong Yang, Min Seo Kim, Xinyu Zhu, Md Mesbah Uddin, Tetsushi Nakao, So Mi Jemma Cho, Satoshi Koyama, Tingfeng Xu, Laurens F. Reeskamp, Rufan Zhang, Zhaoqi Liu, A Yunga, Paul S. de Vries, Ramachandran S. Vasan, Eric Boerwinkle, Alanna C. Morrison, Bruce M. Psaty, Russell P. Tracy, Susan R. Heckbert, Michael H. Cho, Jeong H Yun, Nicholette D. Palmer, Donald W. Bowden, Joanne M. Murabito, Daniel Levy, Nancy L. Heard-Costa, George T. O’Connor, Lewis C. Becker, Brian G. Kral, Lisa R. Yanek, Laura M. Raffield, Bertha Hidalgo, Jerome I. Rotter, Stephen S. Rich, Kent D. Taylor, Wendy S. Post, Charles Kooperberg, Alexander P. Reiner, Braxton D. Mitchell, Sharon L.R. Kardia, Jennifer A. Smith, Patricia A. Peyser, Lawrence F. Bielak, Dong Keon Yon, Hong-Hee Won, Donna K. Arnett, Albert V. Smith, Stacey B. Gabriel, Patrick T. Ellinor, NHLBI Trans-Omics for Precision Medicine (TOPMed) Consortium, Pradeep Natarajan, Minxian Wang, Akl C. Fahed

## Abstract

Multiple germline and somatic genomic factors are associated with risk of coronary artery disease (CAD), but there is no single measure of risk that integrates all information from a DNA sample, limiting clinical use of genomic information. To address this gap, we developed an integrated genomic model (IGM), analogous to a clinical risk calculator that combines various clinical risk factors into a unified risk estimate. The IGM includes six genetic drivers for CAD, including germline factors (familial hypercholesterolemia [FH] variants, CAD polygenic risk score [PRS], proteome PRS, metabolome PRS) and somatic factors (clonal hematopoiesis of indeterminate potential [CHIP], and leukocyte telomere length [LTL]). We evaluated the IGM on CAD risk prediction in the UK Biobank (N=391,536), and validated it in the Trans-Omics for Precision Medicine (TOPMed) program (N=34,177). The 10-year CAD risk based on the IGM profile ranged from 1.1% to 15.5% in the UK Biobank and from 3.8% to 33.0% in TOPMed, with a more pronounced gradient in males than females. IGM captured the cumulative effect of multiple genetic drivers, identifying individuals at high risk for CAD despite lacking obvious high risk genetic factors, or individuals at low risk for CAD despite having known genetic risk variants such as FH and CHIP. The IGM had the highest performance in younger individuals (C-statistic 0.805 [95% CI, 0.699-0.913] for age ≤ 45 years). In middle age, IGM augmented the performance of the Pooled Cohort Equations (PCE), a clinical risk calculator for CAD. Adding IGM to PCE resulted in a continuous net reclassification index of 33.45% (95% CI, 32.11%-34.76%). We present the first model that integrates all currently available information from a single “DNA biopsy” to translate complex genetic information into a single risk estimate.

## Introduction

The early identification of individuals at high risk for coronary artery disease (CAD) is a fundamental strategy in preventing disease which remains the number one cause of mortality and morbidity.^1^ Utilizing DNA information for CAD risk prediction has gained traction due to its ability to identify early-onset cases, predict risk earlier in life, and augment the performance of existing clinical risk measures.^2,3^ Significant progress has been made in understanding germline genomic risk drivers of CAD. Both monogenic drivers of risk such as pathogenic variants in familial hypercholesterolemia (FH)-related genes (*LDLR*, *APOB,* and *PCSK9*) and polygenic risk scores (PRS) have shown promise in CAD risk stratification, individually and in combination.^4,5^ Moreover, age-related somatic mutations have been associated with increased CAD risk, attributed to clonal hematopoiesis of indeterminate potential (CHIP) and shortened leukocyte telomere length (LTL).^6,7^ Even though germline and somatic genomic variations shape CAD risk, they remain to be studied in aggregate. A single model leveraging all available information from a single DNA biopsy could have the potential to improve risk prediction of CAD.

A comprehensive genomic risk model for CAD would integrate risk from early-life germline mutations with risk from somatic mutations occurring later in life. We previously demonstrated the interplay between monogenic and polygenic risk, highlighting that polygenic background alters the penetrance of FH variants.^4^ Thereafter, Zhao et al. reported that a combination of germline and somatic mutations augments the risk of CAD, as evidenced by the interaction between PRS and CHIP.^8^ This mounting evidence underscores that an ensemble model that integrates all known genetic drivers and their interactions might improve genomic risk prediction of CAD.

As genomic medicine moves towards clinical adoption, it might be beneficial that a single measure of risk is communicated using the entirety of the data points available from an individual’s genome. Mixed information about risk is poised to confuse people unless integrated into a single number that is actionable. This concept has long been established in clinical risk prediction. For example, a single 10-year cardiovascular risk is provided by integrating information from factors such as blood pressure, cholesterol levels, smoking, and diabetes, each of which might indicate low or high risk for an individual.^9^ Similarly, in genomic risk prediction, an individual might have a monogenic FH variant, a low polygenic risk score, no CHIP variant, and a short LTL, challenged by how to interpret this complex combination of genetic risk factors. It is only helpful for the individual in this context to understand the summed effect and risk estimate.

We developed an integrated genomic model (IGM) that uses a single score to maximize the precision in CAD risk prediction and enhance clinical translation. We demonstrated the accuracy, calibration, and added value of this all-at-once model using half a million people from the UK Biobank, and validated its performance in studies contributing to the Trans-Omics for Precision Medicine (TOPMed) program.

## Results

### Developing an Integrated Genomic Risk Model for CAD

We used the UK Biobank as a discovery cohort, including 391,536 individuals (mean [SD] age, 56.5 [8.1] years; 53.8% women) including 28,346 (7.2%) participants who developed CAD over a median follow-up of 12.3 (interquartile range [IQR], 1.6) years. In the UK Biobank, 94.1% of participants were White (n = 368,296), with smaller proportions identifying as Asian (2.3%, n = 9,106), Black (1.6%, n = 6,237), and other ancestry (2.0%, n = 7,897). In contrast, TOPMed showed greater diversity, with 68.3% White (n = 23,333), 25.9% Black (n = 8,858), 2.2% Asian (n = 737), and 3.7% Other ancestry (n = 1,250) (Supplementary Table 1). Whole-genome sequencing data was used to curate and compute features of risk previously shown to be associated with CAD. Specifically, we included two somatic features – CHIP and LTL– and four germline features – FH variants, CAD PRS, a proteome PRS (ProPRS) and a metabolome PRS (MetPRS). Of note, ProPRS and MetPRS were constructed using a series of genetic proxies for protein and metabolite levels derived from the atlas of genetic scores that predict multi-omics traits,^10^ rather than relying on actual measurement of protein and metabolite levels. Using this feature matrix, we developed a somatic risk score, a germline risk score, and an integrated genomic model (IGM) (Fig. 1).

**Fig. 1.**
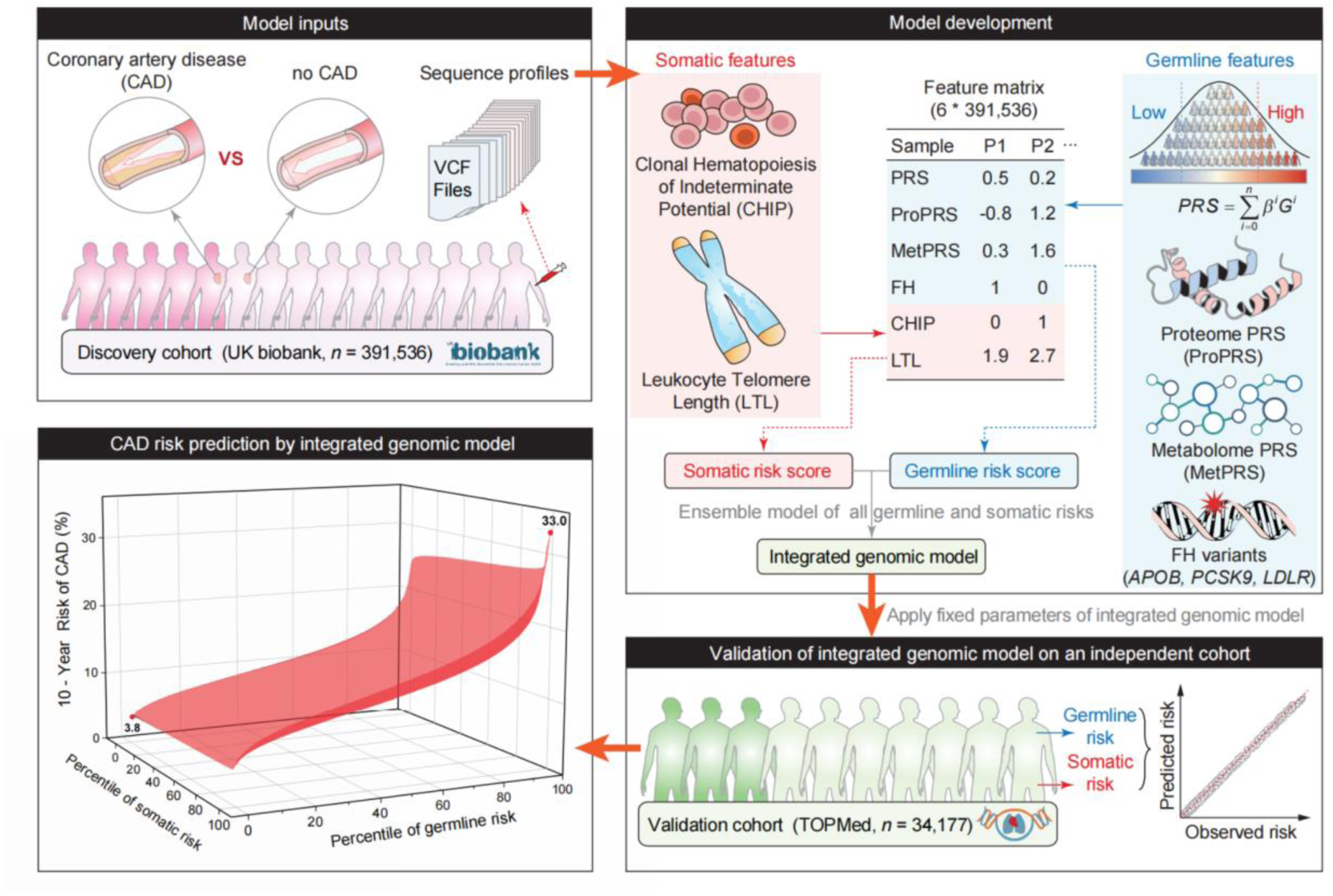
Overview of development and validation of integrated genomic model (IGM). Based on sequencing data from UK Biobank (N=391,536), we curated 6 genomic features that are associated with the risk of CAD. Scores of somatic and germline risks were ensembled to construct IGM, which was then validated in the TOPMed cohort (N=34,177).

We then validated the IGM in 34,177 individuals from the TOPMed program (mean [SD] age, 62.6 [10.6] years; 66.0% women) (Supplementary Table 1). Incident CAD events occurred in 3,972 (14.3%) participants who developed CAD over a median follow-up of 10.5 (IQR, 8.6) years. The IGM model provided individual 10-year risk estimates across percentiles of somatic and germline risk (Fig. 1). Further details on baseline characteristics by genetic drivers are presented in Supplementary Tables 2-5.

### Germline and Somatic Genomic Drivers of CAD Risk

We first estimated the individual risk of prevalent and incident CAD imparted by each of the two somatic and four germline drivers. When evaluating the association of the germline drivers with prevalent CAD in the UK Biobank, FH variant carriers had a three-fold increase in risk – odds ratio (OR) of 3.08 (95% confidence interval [CI], 2.46-3.85; p < 0.001). The CAD PRS (OR per standard deviation (SD), 2.15; 95% CI, 2.11-2.19; p < 0.001), MetPRS (OR per SD, 1.27; 95% CI, 1.25-1.29; p < 0.001), and ProPRS (OR per SD, 1.19; 95% CI, 1.17-1.21; p < 0.001) were also significantly associated with CAD (Fig. 2A). We combined these four germline risk factors into a single predictor called GermRisk. The OR per SD for GermRisk was 2.16 (95% CI, 2.12-2.20; p < 0.001), and individuals in the top quintile of GermRisk had 3.6-fold increase in risk compared to everyone else (95% CI, 3.47-3.73; p < 0.001) (Supplementary Table 6). The effect sizes of genetic drivers for prevalent CAD in TOPMed were overall consistent with those of UK Biobank (Fig. 2B; Supplementary Table 7).

**Fig. 2.**
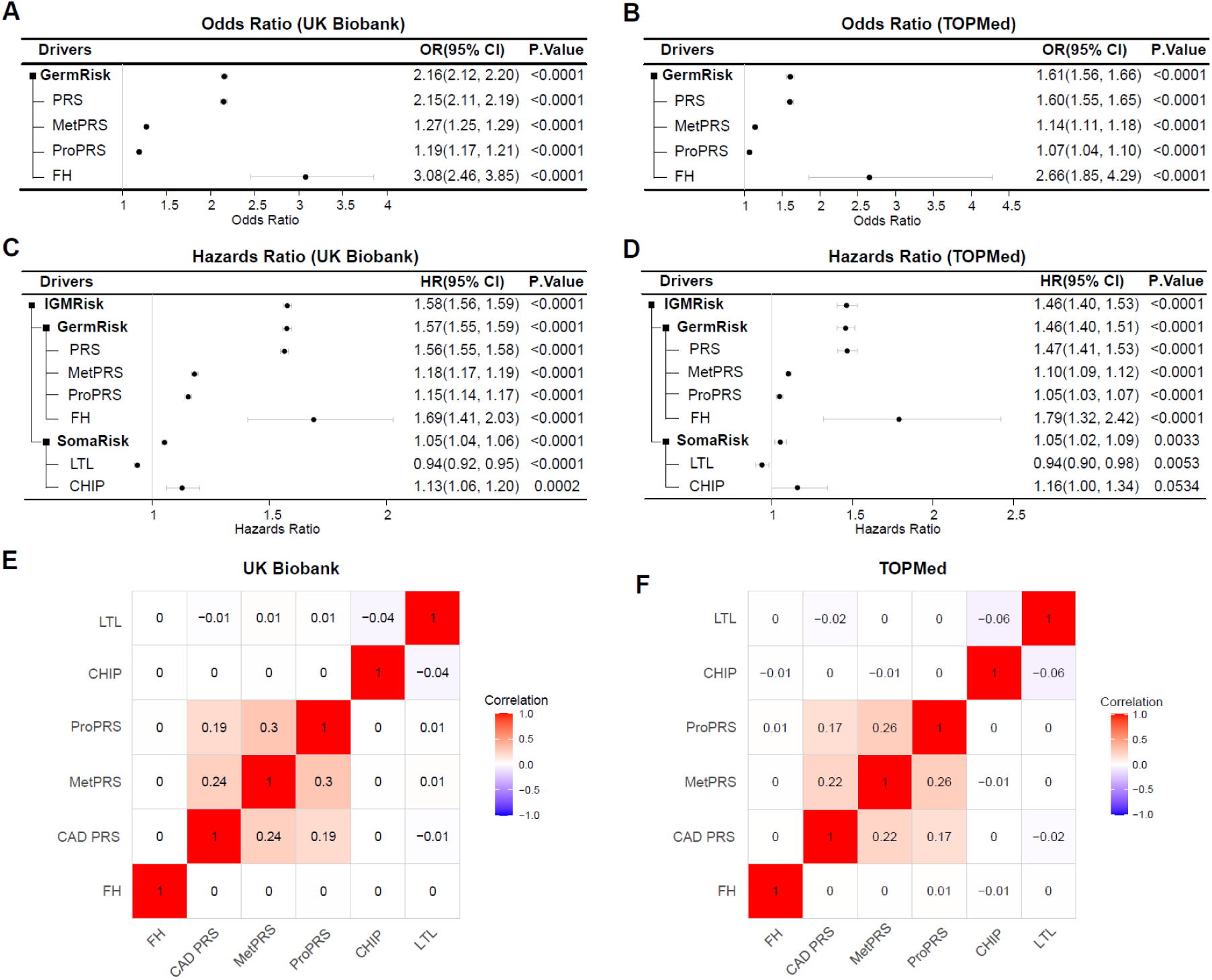
Effect size of genomic drivers in UK Biobank and TOPMed. Effect sizes based on prevalent coronary artery disease (CAD) in UK Biobank (**A**) and TOPMed (**B**), and incident CAD in UK Biobank (**C**) and TOPMed (**D**) are presented. Odds ratio and hazard ratio per standard deviation are shown for all continuous measures (PRS, MetPRS, ProPRS, LTL, GermRisk, SomaRisk, IGMRisk). Odds ratio and hazard ratio per carrier status are shown for FH and CHIP. Estimates are derived from logistic regression (panels A and B) or Cox proportional hazards model (panels C and D) with sex, recruitment age, and the first 10 principal components of genetic ancestry as covariates. GermRisk is a weighted combination of four genetic drivers (PRS, MetPRS, ProPRS, and FH) as a single predictor, with weights estimated by a logistic regression model. SomaRisk is a weighted combination of two somatic drivers (CHIP and LTL) as a single predictor, with weights estimated by a Cox proportional hazards model. IGMRisk is a combination of GermRisk and SomaRisk, weighted from a Cox proportional hazards regression model estimation. Correlations among the six genetic drivers are shown for the UK Biobank (**E**) and TOPMed (**F**). CI, confidence interval; PRS, polygenic risk score (PRS) for CAD; MetPRS, metabolome PRS; ProPRS, proteome PRS; LTL, leukocyte telomere length; FH, familial hypercholesterolemia variants; CHIP, clonal hematopoiesis of indeterminate potential.

We then evaluated the association of genomic drivers with incident CAD in the UK Biobank, and demonstrated strong associations of each of the germline and somatic drivers with incident CAD (Fig. 2C; Supplementary Table 8). For germline drivers, the HR for FH carriers was 1.69 (95% CI 1.41-2.03, p < 0.001) and HRs per SD for CAD PRS, MetPRS, and ProPRS were 1.56 (95% CI 1.55-1.58, p < 0.001), 1.18 (95% CI 1.17-1.19, p < 0.001), and 1.15 (95% CI 1.14-1.17, p < 0.001), respectively. For somatic drivers, CHIP and LTL were associated with CAD with HR of 1.13 (95% CI 1.06-1.20, p < 0.001) and 0.94 (95% CI 0.92-0.95, p < 0.001), respectively. We combined these two somatic risk factors into a single predictor called SomaRisk, similar to GermRisk. Both GermRisk and SomaRisk demonstrated a strong association with incident CAD – HR per SD of 1.57 (95% CI 1.55-1.59, p < 0.001) and 1.05 (95% CI 1.04-1.06, p < 0.001), respectively (Fig. 2C; Supplementary Table 8). The effect sizes of genetic drivers for incident CAD in TOPMed were mostly consistent with those of UK Biobank (Fig. 2D; Supplementary Table 9).

### Integrated Genomic Model to Predict CAD Risk

We assessed pairwise correlations between six genetic variables from the UK Biobank and TOPMed studies using Pearson correlation coefficients to evaluate multicollinearity before combining them in a Cox proportional hazards model. This allowed us to gauge overlapping signals, particularly between CAD PRS, MetPRS, and ProPRS. The correlations among six drivers were weak (≤ 0.3) (Fig. 2E and 2F), reassuring that each driver contributes distinct signals.

To obtain a comprehensive assessment of a person’s CAD risk, we used an integrated genomic model (IGM) to quantify the risk from both germline and somatic drivers – a combined predictor of GermRisk and SomaRisk (Supplementary Fig. 1). The IGM risk was significantly associated with the risk of incident CAD (HR per SD, 1.58; 95% CI, 1.56-1.59; p < 0.001), and the effect size was consistent when validated in the TOPMed external data set (HR per SD, 1.46; 95% CI, 1.40-1.53; p < 0.001) (Fig. 2C and 2D; Supplementary Tables 8-9).

Joint modeling of germline and somatic drivers indicated substantial gradients in risk of CAD, according to inherited DNA variants and variation in the rate of LTL shortening and accumulation of somatic variants leading to CHIP. For the sex-combined estimation of the 10-year risk of CAD in UK Biobank, individuals in the lowest germline and somatic risk percentile have a 10-year risk as low as 1.1%, while those in the highest germline and somatic risk percentile have a 10-year risk as high as 15.5% (Fig. 3A). A similar gradient in risk across germline and somatic variation was observed in TOPMed, ranging from 3.8% to 33.0% (Fig. 3B). For a sex-stratified analysis in the UK Biobank, male individuals had a 10-year risk that ranged from 1.8% to 23.0% across the germline and somatic risk spectrum. This was about 2.3 times higher than the risk spectrum for females, which spanned from 0.7% to 10.3% (Fig. 3C). Large gradients in 10-year risk were consistently observed in the TOPMed for the sex-stratified analysis, with males ranging from 4.8% to 39.9% and females ranging from 3.0% and 27.0% (Fig. 3D).

**Fig. 3.**
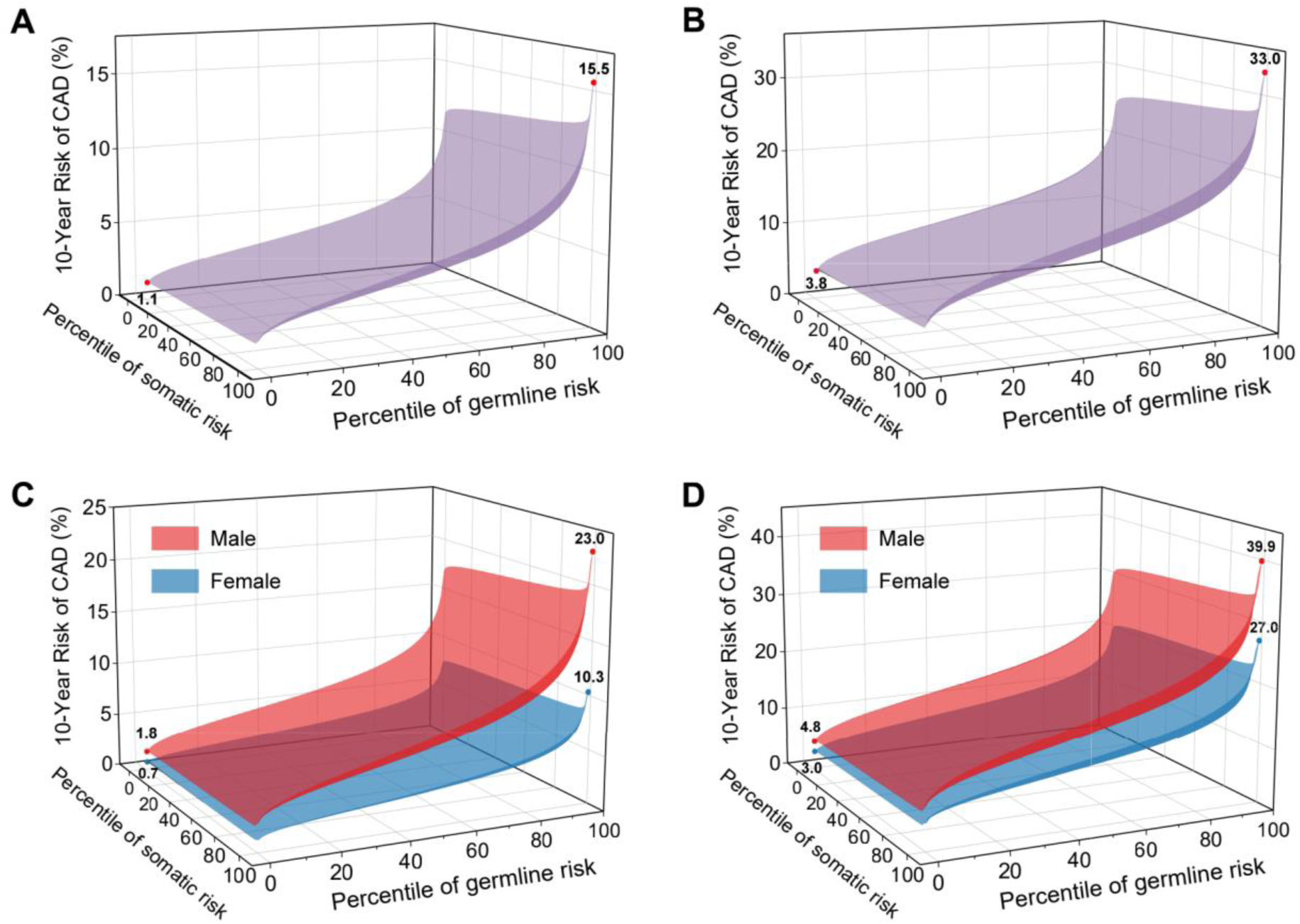
Ten-year risk of CAD as a function of somatic and germline risk from the integrated model. Ten-year risk of CAD among all participants from UK Biobank (**A**) and TOPMed (**B**), and sex-stratified 10-year risks of CAD in UK Biobank (**C**) and TOPMed (**D**) are presented.

### Heterogeneity of Genomic Risk Profiles Captured by the Integrated Genomic Model

The IGM effectively captured a range of genetic risk combinations for CAD, identifying high risk groups (top 20% overall risk) with diverse genetic profiles in the UK Biobank (Fig. 4A). High risk IGM group included individuals at high risk in both germline and somatic factors (20.6%), those at high risk for one of the factors (78.6%), and a small proportion with moderately elevated, yet sub-threshold, risks for both factors (0.8%) (Fig. 4A). Individuals at high risk for both germline and somatic factors had the highest 10-year risk (8.8%, 95% CI 8.43-9.19%) within the high risk IGM group (Supplementary Table 10). As expected, the high risk group identified by the IGM had a larger number of genetic risk drivers compared to the low risk group. For example, 63.9% had two or more genetic drivers in the high risk group, compared to only 3.4 % in the low risk group in the UK Biobank (Fig. 4B). Notably, people at low IGM risk were not without genetic risk drivers, and a non-negligible proportion of 29.3% had one or more genetic risk drivers (Fig. 4B and Supplementary Table 11); however, mitigating effects from other genetic variants (e.g., low CAD PRS) seem to offset the overall risk, thus classifying these individuals as low risk. Similar proportions of breakdown and distribution patterns were observed in TOPMed (Fig. 4C, 4D and Supplementary Table 11).

**Fig. 4.**
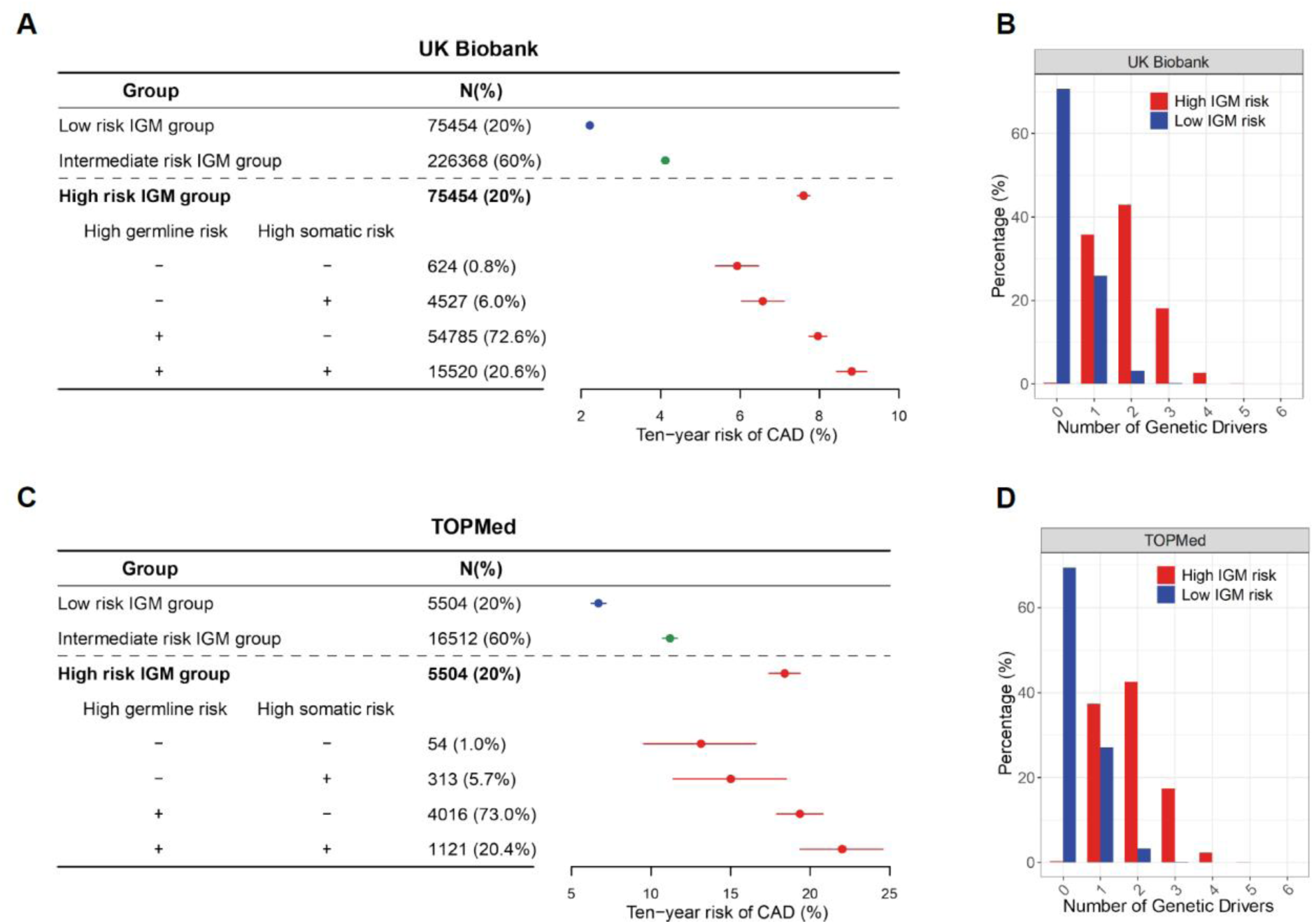
The integrated genomic model (IGM) captures participants with diverse genetic risk profiles contributing to risk in the UK Biobank (**A, B**) and TOPMed (**C, D**). (**A**) and (**C**), The ten-year risk of CAD was estimated for each IGM risk group in the UK Biobank and TOPMed dataset. Categories were defined as low risk (bottom 20%), intermediate risk (middle 60%), and high risk group (top 20%). In the high risk group, the genetic profile was further partitioned by the status of carrying a high germline risk or high somatic risk. The high germline risk was defined as the top 20% with a composite risk estimated from four germline genetic risk drivers (FH, PRS, MetPRS, and ProPRS). The high somatic risk was defined as the top 20% with a composite risk estimated from two somatic risk drivers (CHIP and LTL). (**B**) and (**D**), the genetic risk profiles for the IGM low risk (bottom 20%) and high risk (top 20%) groups, respectively. The six drivers are FH, PRS, MetPRS, ProPRS, CHIP, and LTL. The continuous variables were binarized, with individuals in the top 20% treated as carriers, except for LTL (the bottom 20% were treated as carriers). PRS, polygenic risk score (PRS) for CAD; MetPRS, metabolome PRS; ProPRS, proteome PRS; LTL, leukocyte telomere length; FH, familial hypercholesterolemia variants; CHIP, clonal hematopoiesis of indeterminate potential.

### Integrated Genomic Model and Clinical Risk

The American College of Cardiology/American Heart Association, Pooled Cohort Equations (PCE) are a guideline-recommended clinical-risk calculator that uses clinical risk factors to identify high risk people for initiation of preventive treatments (i.e., statin).^11^ Within each of the guideline-defined strata of the PCE risk, the IGM score was a strong predictor of coronary artery disease events and showed a consistent risk gradient by IGM score category (Fig. 5A, B). Among participants at high PCE risk (> 20% 10-year risk), the 10-year CAD event rates were 5.4%, 9.5%, and 16.4%, for low, intermediate, and high IGM risk groups (Fig. 5A). Remarkably, a 10-year CAD risk threshold of 7.5% for initiating statin therapy as guideline recommended was reached even among individuals in the borderline (5.0%-7.4% 10-year risk) and intermediate (7.5%-19.9% 10-year risk) PCE categories when stratified by IGM percentiles (Fig. 5B) – an IGM score higher than 95th and 72nd percentile, respectively for borderline and intermediate risk (Fig. 5B). A close match of the model predicted and actual observed 10-year disease risk shows the models were well calibrated (Supplementary Fig. 2). When disaggregated to individual genetic risk drivers, the FH variants and CAD PRS most prominently re-stratified the CAD risk across PCE categories, but other risk factors also have significant stratification ability (Supplementary Fig. 3).

**Fig. 5.**
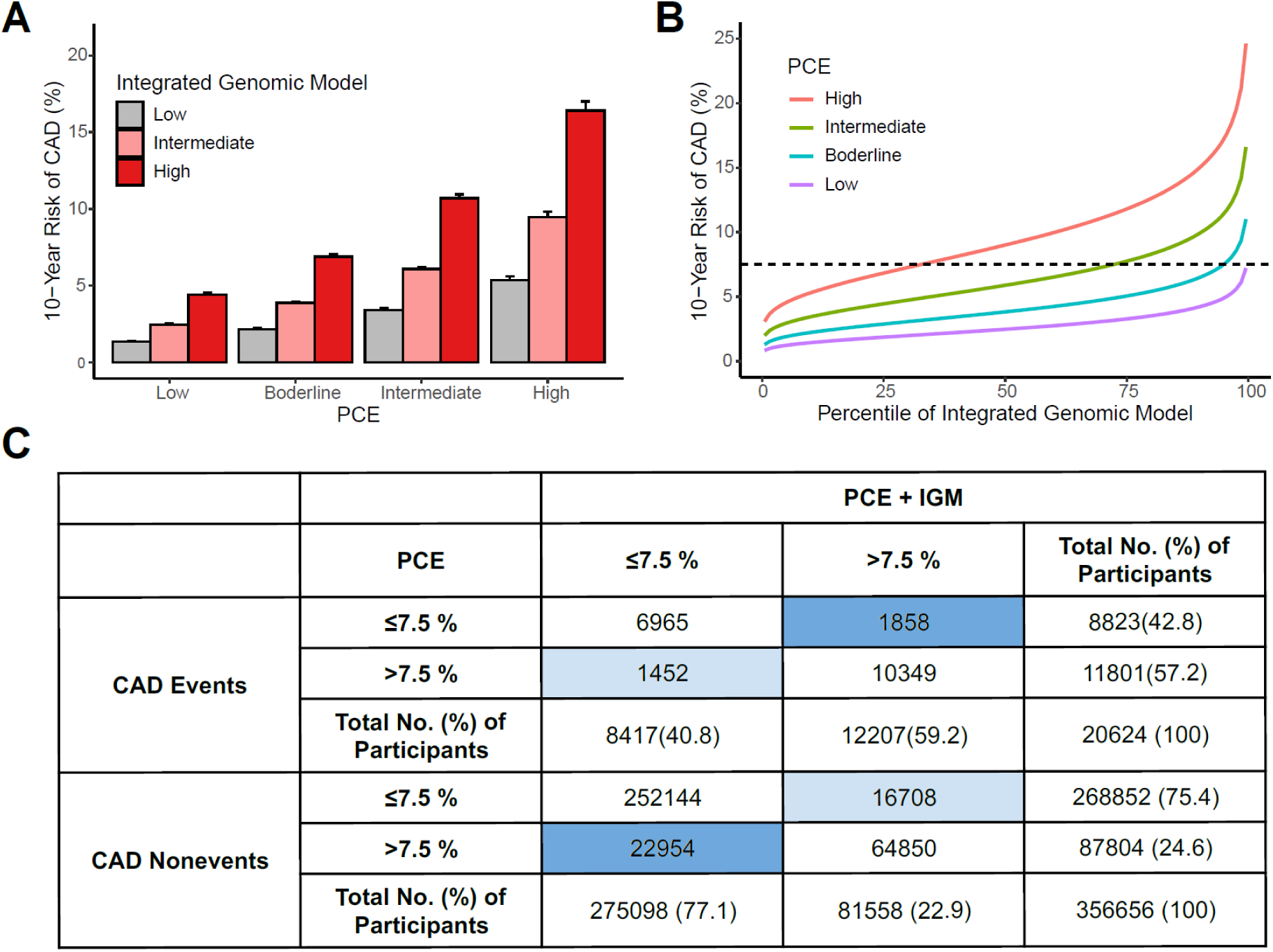
Stratification (**A**, **B**) and reclassification (**C**) of 10-year predicted CAD risk based on IGM. **A**, stratification by IGM risk within PCE risk stratum. **B**, Predicted 10-year CAD risk gradient by genetic risk percentile. **C**, Reclassification of 10-year predicted CAD risk–columns and rows indicate categories of 10-year predicted risk, with the number of individuals in each risk category, the number of samples correctly reclassified and wrongly reclassified are in dark and light blue, respectively. A continuous net reclassification index was 33.45% (95% CI, 32.11%-34.76%). IGM categories were defined as low risk (bottom 20%), intermediate risk (middle 60%), and high risk group (top 20%), respectively. PCE categories were defined as low (estimated risk less than 5%), borderline (risk between 5% to 7.5%), intermediate (risk between 7.5% and 20%), and high (risk greater than 20%), respectively. IGM: integrated genomic model; PCE, pooled cohort equation.

By combining the conventional clinical risk of PCE with the genetic risk of IGM, the model showed the most potent risk stratification ability. When 20,624 individuals who experienced CAD events in the UK Biobank were used to determine reclassification by adding IGM to PCE compared to PCE alone, 1,858 were correctly classified at a higher risk, while 1,452 were incorrectly placed at a lower risk (Fig. 5C), leading to a net proportion of accurate reclassifications for events is 1.97% (406/20,624). For nonevents, 22,954 individuals were correctly down-classified, and 16,708 were incorrectly up-classified, leading to a net reclassification proportion of 1.75% (6,246/356,656) for nonevents. The overall NRI combines the events and nonevents, resulting in 3.72% (95% CI, 3.15%-4.26%). Continuous NRI was 33.45% (95% CI, 32.11%-34.76%). As shown in the full stratification table, there was a lower event rate in the low risk category for the combined model than in the PCE-alone model (1.9% versus 2.3%), however, a higher event rate was observed for the high risk category (27.1% versus 23.5%), demonstrating the prominent risk stratification ability of the IGM model with guideline-recommended PCE risk estimator (Supplementary Table 12).

Finally, we evaluated the discrimination of PCE, IGM, and combined model compared to a baseline model with age, sex and genetic ancestry. The C-statistic for the base model was 0.701 (95% CI, 0.698-0.704), PCE was 0.725 (95% CI 0.722-0.728), and the combination of base and IGM model was 0.734 (95% CI 0.731–0.737), respectively, in UK Biobank (Supplementary Fig. 4A; Supplementary Table 13). The performance was highest when combining PCE and IGM (C-statistic, 0.750; 95% CI, 0.747-0.753). In the TOPMed data, the discrimination with IGM was further improved when limiting prediction to younger individuals (aged ≤ 45 years) (C-statistic 0.805, 95% CI, 0.699-0.913)(Supplementary Fig. 4B and Supplementary Table 13).

## Discussion

We developed an integrated genomic model for CAD prediction that combines multiple known germline and somatic risk drivers that can be measured using a single DNA biopsy. The model demonstrated value in improving the precision of risk estimation and capturing people at risk due to diverse genetic risk profiles. Based on the IGM, the 10-year CAD risk varied from 1.1% to 15.5% among UK Biobank participants and 3.8% to 33.0% in TOPMed study participants, with a more pronounced gradient in males than females for both cohorts. The integrated genomic score showed a high discrimination when combined with clinical risk score or used in younger age groups. The addition of the IGM to the clinical risk model resulted in a continuous net reclassification index of 33.45%. The integrated genomic model captured cumulative and comprehensive effects of multiple genetic drivers, identifying high risk individuals who may otherwise be overlooked when using conventional risk models relying on a single genomic driver (i.e., PRS or FH).

Conventional genomic risk models for CAD that focused on monogenic (FH) or polygenic (PRS) drivers were limited in their uniaxial approach, falling short of encompassing the wide range of genetic combinations present in real-world populations.^12^ For instance, individuals carrying FH and CHIP variants might still have a low overall risk of CAD if their effects are offset by protective PRS and low MetPRS. Other individuals may present with different combinations of risk and protective genetic factors that collectively indicate a high CAD risk. Such dynamics complicate CAD risk assessment using standard genomic approaches that rely on stratification by a single genetic driver. Our IGM captured such dynamics by integrating all known genomic risk drivers for CAD, including germline, somatic, and predicted proteomic/metabolomic drivers in a single model, without compromising the performance. We successfully demonstrated that IGM captures cumulative and comprehensive effects of multiple genetic drivers, identifying high risk individuals who do not have obvious germline and somatic risk but whose aggregate genetic risk escalates to a high overall risk (Fig. 4A, 4B). Conversely, a subset of individuals classified in the low risk group by IGM possessed one or more high risk genetic drivers (Fig. 4C), indicating other drivers may confer protective benefits and thus reduce the overall risk. Such a dynamic profile of personal genomics should be considered to fully achieve the goals of precision prevention and personalized care.

The high risk group identified by the IGM exhibited an impressive diversity in their genetic profiles and combinations. In the high risk group identified by IGM, 20.9% and 19.8% of individuals carried a high risk classification for more than 3 genetic drivers while 36.1% and 37.6% had a high risk for only 1 or 0 drivers in UK Biobank and TOPMed studies, respectively (Supplementary Table 11). Interestingly, in the low risk group by IGM (bottom 20%), there were 32 and 593 UK Biobank participants carrying FH and CHIP variants respectively, suggesting that a single genetic factor does not necessarily dictate the overall risk for the disease (Supplementary Table 14). Our observation unveiled a cumulative pattern where the protective level of one or a group of drivers can offset the risk posed by others. The multidimensional nature of our model might facilitate a nuanced approach to risk stratification and draw clinical attention to at-risk individuals who would have otherwise been overlooked by conventional genomic models that are not designed to capture diversity of the genetic pool.

The integrated genomic model based on comprehensive DNA information is a strong predictor of CAD in young adults enabling primordial prevention prior to the onset of clinical risk factors. While IGM had modestly higher discriminative capacity for incident CAD compared with the clinical risk score, its predictive accuracy was significantly higher in younger individuals (aged ≤45 years) in the TOPMed program studies (C-statistic 0.805) (Supplementary Table 13). Our findings imply that the use of genetic information to predict future CAD risk might be more profound for young adults, consistent with previous findings.^13,14^ In contrast to clinical risk models, the IGM is available even before clinical risk factors manifest, providing an additional benefit to young adults who often remain undetected on the radar of traditional assessments.

The highest prediction was achieved by combining IGM with clinical risk score, highlighting the value of genetics in complementing clinical risk prediction. The IGM enabled risk stratification for CAD within each clinical risk stratum, enhancing the identification of individuals requiring targeted clinical interventions. Low genetic risk individuals in the high PCE group demonstrated equivalent 10-year CAD risk with average genetic risk individuals in the intermediate PCE group, and a consistent downward trend was observed across all clinical risk strata (Fig. 5A). Conversely, an upward trend was observed for individuals with high genetic risk identified by IGM. Current guidelines recommend initiating statin therapy for individuals in the intermediate PCE category, defined as a 10-year CAD risk of 7.5% or higher.^15^ However, our findings indicate that individuals in the borderline PCE category who have a high genetic risk may also warrant targeted interventions, as their 10-year CAD risk is comparable (Fig. 5A). It is noteworthy that individuals at the 33rd, 72nd, and 95th percentiles of integrated genomic risk all exhibited an equivalent 10-year CAD risk of 7.5% (Fig. 5B), despite being categorized in high, intermediate, and borderline PCE groups, respectively. This further indicated that existing clinical-focused models might not adequately encompass the multifaceted nature of CAD risk, warranting the consideration of the interplay between genetic and clinical factors in risk evaluations.

While monogenic and polygenic drivers of risk have become well-established, emerging models based on proteomic data are now being developed to predict cardiovascular risk, expanding beyond traditional clinical and genomic models.^16,17^ Helgason et al. recently developed a protein risk score based on 4,963 plasma proteins from 13,540 Icelanders and demonstrated reliable predictability for major cardiovascular risk.^16^ Nevertheless, implementation of proteome measurement in clinical practice remains challenging due to the cost and feasibility constraints. To make the most of proteomic and metabolomic insights in settings without their direct measurements, we developed proteome and metabolome PRSs, leveraging genetically-predicted protein and metabolite levels instead of actual serum protein levels based on the genetic score atlas for multi-omics traits.^10^ We calculated genetic score for 2,692 proteins and 876 metabolites levels, with 124 proteins and 142 metabolites comprising the final ProPRS and MetPRS models after Lasso penalty was applied, respectively (Supplementary Fig. 5; Supplementary Tables 15-16). Although some correlation was present among the MetPRS, ProPRS and CAD PRS, the magnitude was weak (≤ 0.3) (Fig. 2), implicating that each driver contributes a distinct, non-overlapping signal. To the best of our knowledge, we have introduced the first genetically-predicted protein and metabolite risk scores for CAD risk prediction, which are readily obtainable through standard low-cost DNA microarray or sequencing and thereby more cost-effective than serum protein measurements. Our purpose was to make the most out of a single DNA biopsy, which is becoming increasingly feasible through adoption of genomic medicine and large biobanking efforts.

This study has several limitations. First, we have not provided ancestry-specific results given the majority composition with European and smaller sample size for non-European ancestries. However, to promote inclusion and equity, we used multi-ancestry cohorts from UK Biobank and TOPMed in the primary analysis, as well as multi-ancestry PRS that has been demonstrated to perform well for both European and non-European ancestries.^12^ Second, this study evaluated CAD as the primary outcome whereas PCE was developed to predict cardiovascular disease which includes CAD and stroke. Nevertheless, previous studies have shown that the PCE is effective in predicting CAD.^18,19^ Third, the CAD PRS used in this study included low frequency and common variants, but did not incorporate rare high-impact genetic variants associated with CAD risk. However, based on whole genome sequencing data from half a million populations, we separately identified somatic mutations and curated significant rare variants such as those linked to FH to develop a comprehensive genomic model. This approach allowed us to comprehensively capture the diverse spectrum of genetic variant frequencies linked to CAD. Fourth, baseline CAD risk is different in the UK Biobank and TOPMed because UK Biobank consists of healthier individuals compared to TOPMed. The incidence of CAD was 7.2% and 12.7% respectively for UK Biobank and TOPMed (Supplementary Table 1), and this was reflected in the risk gradient captured by IGM the 10-year CAD risk ranged from 1.1% to 15.5% among UK Biobank participants, and 3.8% to 33.0% in TOPMed participants (Fig. 3).

## Conclusion

We integrated all currently available information from a single “DNA biopsy” to translate complex genetic information into a single risk estimate. The IGM powerfully stratifies CAD risk in young individuals and complements clinical risk prediction in middle-aged individuals. Because the model considered the contributions of multiple genomic drivers for every individual, it was able to identify high risk individuals who may otherwise be overlooked when using conventional risk models relying on a single genomic driver. Our model holds the promise to transform CAD risk assessment strategies in an emerging ’genome-first’ healthcare framework, where genomic information becomes readily accessible and a fundamental part of patient care. Moreover, the framework we propose could be extended to other diseases known to have multiple genomic risk drivers.

## Methods

### Dataset and Quality Control

Access to the UK Biobank data was approved with application ID 89885. Samples with discordance between self-reported sex (Field 31) and genetically inferred sex (Field 22001) were removed. Additionally, samples with individual-level genotype missing rates (Field 22005) greater than 5%, outliers for heterozygosity or missing rate (Field 22027), or sex chromosome aneuploidy (Field 22019) were excluded. To remove close relatives in the samples, we excluded one of the samples whose pairwise kinship value is greater than or equal to 0.0884 (threshold of the second-degree close relatives^20^) but also tried to keep as many samples as possible. Finally, 391,536 individuals were included in the final analysis (Supplementary Fig. 6A).

A total of 80,588 participants from the NHLBI’s TOPMed program with available whole genome sequencing data were considered. These studies primarily consist of observational cohorts that have been described in detail previously.^6^ Among 80,588 participants, we excluded 49 samples with conflicting sex information, 1 sample without principal components, and 17,237 samples with excess kinship, defined as a second-degree relationship or closer, indicated by a KING coefficient greater than 0.0884. Finally, 34,177 participants remained for analysis after excluding an additional 29,124 participants without CAD phenotype (Supplementary Fig. 6B). Cohorts contributed to this population are Amish, Atherosclerosis Risk in Communities Study[ARIC], Cardiovascular Health Study[CHS], Genetic epidemiology of COPD[COPDGene], Diabetes Heart Study[DHS], Framingham Heart Study[FHS], Genetic Study of Atherosclerosis Risk[GeneSTAR], Genetic Epidemiology Network of Arteriopathy[GENOA], Jackson Heart Study[JHS], Multi-Ethnic Study of Atherosclerosis[MESA], and Women’s Health Initiative[WHI].

### Curation of FH, CHIP, and LTL

To determine the carrier status of familial hypercholesterolemia (FH) for samples with available whole-exome sequencing data, a combination of variant selection criteria was applied: 1) Variants from previous publications which were manually curated by clinical geneticists;^4,21–23^ 2) Variants in *LDLR, APOB* and *PCSK9* from the ClinVar database (downloaded February 27th, 2023, GRCh38) annotated as pathogenic, likely pathogenic, or pathogenic/likely pathogenic without conflicts. For *APOB* and *PCSK9* gene, only variants associated with hypercholesterolemia but not hypobetalipoproteinemia were included; 3) Variants in *LDLR* annotated as high-confidence loss-of-function by the VEP Loss-Of-Function Transcript Effect Estimator (LOFTEE) plugin were also included.^24,25^ Only variants with a net positive association with LDL cholesterol level accessed by an iterative conditional regression analysis were included in the final variant list for subsequent analysis (Supplementary Table 17).^26^

The carrier status of clonal hematopoiesis of indeterminate potential (CHIP) was detected following a similar procedure described in Yu Zhi et al. and others.^27–29^ Specifically, carrier status was determined by carrying CHIP variants from one of the genes *TET2, ASXL1, JAK2, PPM1D, TP53, SRSF2, and SF3B1*. Leukocyte telomere length (LTL) was log-transformed to obtain a normal distribution and then Z-standardized using the distribution of all individuals with a telomere length measurement (Field 22192). Details of processing were described in V. Codd et al.^30^ Unless otherwise specified, genetic drivers have been curated comparably for UK Biobank and TOPMed. More details on the curation of CHIP and LTL for TOPMed are described elsewhere.^6^

### CAD Polygenic Risk Score, Metabolome Polygenic Risk Score, and Proteome Polygenic Risk Score

In the UK Biobank, CAD PRS was calculated using the imputed genotype data and variant weights from the multi-ancestry and multi-trait polygenic risk score for CAD described in A. P. Patel et al.,^12^ implemented with PLINK2.^31^ Proxy Risk scores for Metabolome (Metabolon and Nightingale) and Proteome (Somalogic and Olink) were calculated with the imputed genotype data and model weight files downloaded from OMICSPRED resource which derived from the INTERVAL study cohort (https://www.omicspred.org/Scores/Somalogic/INTERVAL),^10^ with 726 (Metabolon), 141 (Nightingale), 2,384 (Somalogic), and 308 (Olink) scores, respectively. We randomly sampled 200,000 individuals from UK Biobank as the training set, and a lasso penalty was applied to the Cox proportional hazards regression model with age, sex, and the top 10 principal components (PCs) as covariates and incident CAD as an outcome, similar to the process described elsewhere.^16^ Five-fold cross-validation was employed to select the optimal penalization strength for hyperparameters. Two independent models were trained for the prediction of CAD risk using proxy scores of metabolome (genetic risk scores for 867 serum metabolites) and proteome (genetic risk scores for 2,692 serum proteins). With weights determined by the cross-validation procedures, the weighted proxy risk scores for Metabolome (MetPRS) and Proteome (ProPRS) were then calculated for all samples. The number of genetically predicted metabolome and proteome scores retained in the MetPRS and ProPRS lasso models were 142 and 124, respectively (Supplementary Tables 15-16).

### CAD Definition

In the UK Biobank, CAD was defined based on self-report at enrollment, hospitalization records, or death registry records as previously described (Supplementary Table 18).^4^ In the TOPMed studies, CAD was defined as ischemic heart disease events, including myocardial infarction and coronary revascularization.^6^ Incident CAD cases were defined as those diagnosed after recruitment. The survival year was defined as the years between recruitment and diagnosis for incident CAD cases, or between the time of the recruitment and the last censoring for controls.

### Covariates and Adjustment

Untreated blood pressure was estimated by adjusting the raw value for anti-hypertensive medication intake by adding 15 mmHg to the systolic blood pressure and 10 mmHg to the diastolic blood pressure as previously described.^32,33^ Untreated lipid levels were estimated by adjusting the raw lab-tested value according to lipid-lowering medication intake as described previously^12^ and detailed in Supplementary Table 19.

To eliminate potential confounding effects of covariates, PRS, MetPRS, ProPRS, and LTL were regressed on recruitment age, sex, and the first 10 principal components of genetic ancestry (Supplementary Fig. 7). The scaled residuals with mean zero and standard deviation of one were then used in the subsequent analyses.

### Developing the integrated genomic risk model

We evaluated pairwise correlations between six genetic variables from the UK Biobank and TOPMed studies to assess potential multicollinearity before including these variables in a regression model. The correlation matrix was computed using Pearson correlation coefficients. As germline genetic risk drivers (FH, PRS, MetPRS, and ProPRS) remain constant from birth, while CHIP accumulates and LTL shortens with age (somatic risk drivers), we systematically characterized and investigated the joint effects of germline and somatic risk drivers on CAD. Germline risks were combined into a single value termed GermRisk, and somatic risks were combined into a single value termed SomaRisk, the combination weights were estimated by a joint regression model. Specifically, for prevalent CAD, the germline risk drivers (FH, PRS, MetPRS, and ProPRS) were fitted into a logistic regression model, and the coefficients were used as weights to combine the values of the drivers into GermRisk linearly. For incident CAD, the germline risk drivers were fitted to a Cox proportional hazards model, and GermRisk was calculated as a weighted summation of germline variables and their corresponding Cox coefficients, and similar procedures were performed to obtain SomaRisk.

Next, GermRisk and SomaRisk were regressed on recruitment age, sex, and the top 10 PCs, and the residuals were scaled to have zero mean and unit standard deviation, respectively. The standardized residuals of GermRisk and SomaRisk were then fitted to a Cox proportional hazards model. A final predictor, termed IGM (integrated genomic model), was a linear summation of GermRisk and SomaRisk residual according to the Cox coefficient estimation. The weights were fixed and applied to TOPMed data for independent validation. The overall framework for developing and validating the integrated genomic model is shown in Fig. 1.

### Estimating Effect Sizes

To investigate the genomic factors driving the risk of prevalent and incident CAD, a logistic regression model and Cox proportional hazards regression model were employed respectively to estimate the effect sizes for individual genetic drivers. Additionally, PRS, MetPRS, ProPRS, LTL, and the ensembled ones (IGMRisk, GermRisk and SomaRisk) were binarized, with individuals in the top 20% treated as carriers (hPRS, hMetPRS, hProPRS, hIGMRisk, hGermRisk and hSomaRisk) or bottom 20% treated as a carrier (sLTL) for telomere length.

### Interplay Between Genomic Drivers and Clinical Risk Score

To investigate the interplay between genomic risk and PCE, participants in the UK Biobank were divided into four groups based on guideline-defined categories of the PCE – low (estimated risk less than 5%), borderline (risk between 5% to 7.5%), intermediate (risk between 7.5% and 20%), and high (risk greater than 20%).^15^ For the IGM, we divide samples into three risk groups as standard in genomic analyses – high genomic risk (top quintile of the distribution), intermediate genomic risk (middle three quintiles), and low risk (bottom quintile).

### Estimating 10-year risk of CAD

The 10-year risk of CAD was estimated by Cox proportional hazards regression with GermRisk and SomaRisk as predictors and the age, sex, and first 10 principal components of genetic ancestry as covariates. To investigate the stratification capacity of different models, the C-statistic from 1) a base model (sex, age, and first 10 PCs), 2) a log-transformed value of PCE, 3) a base model and IGM, and 4) a base model, IGM and log(PCE), was estimated by a Cox proportional hazards model, respectively.

To evaluate the improvement of prediction by adding the IGM to PCE, the net reclassification improvement was calculated based on a 10-year risk threshold of 7.5% for categorical reclassification and threshold 0 for continuous reclassification as demonstrated elsewhere.^18,19^ Confidence intervals were estimated by 100 times bootstrap. All statistical analyses were done using R v4.2.2 (R Foundation, Vienna, Austria), including the following packages: survival (v3.5-7), survminer (v0.4.9), tableone (v0.13.2), pROC(v1.18.5), nricens(v1.6), rms(v6.7-1) and glmnet (v4.1-8).

## Data availability

All data are made available from the UK Biobank (https://www.ukbiobank.ac.uk/enable-your-research/apply-for-access) to researchers from universities and other institutions with genuine research inquiries following institutional review board and UK Biobank approval. This research was conducted using the UK Biobank resource under Application Number 89885 and approved by Beijing Institute of Genomics review board. The weights of MetPRS and ProPRS are available in the Polygenic Score Catalog (IDs: PGS005093-PGS005094). This paper used the TOPMed whole genome sequencing (WGS) data and cardiovascular disease phenotype data. Genotype and phenotype data are both available in database of Genotypes and Phenotypes (dbGaP). The TOPMed WGS data were from the following eleven study cohorts: Amish, Atherosclerosis Risk in Communities Study (ARIC), Cardiovascular Health Study (CHS), Genetic epidemiology of COPD (COPDGene), Diabetes Heart Study (DHS), Framingham Heart Study (FHS), Genetic Study of Atherosclerosis Risk (GeneSTAR), Genetic Epidemiology Network of Arteriopathy (GENOA), Jackson Heart Study (JHS), Multi-Ethnic Study of Atherosclerosis (MESA), and Women’s Health Initiative (WHI).

## Acknowledgements

Dr. Ellinor is supported by grants from the National Institutes of Health (R01HL092577, 1R01HL157635, 5R01HL139731), from the American Heart Association (18SFRN34110082, 961045) and from the European Union (MAESTRIA 965286). Dr. Natarajan is funded by grants R01HL1427, R01HL148565 R01HL148050, and U01HG011719 from the National Institutes of Health. Dr. Fahed is funded by grants K08HL161448 and R01HL164629 from the National Institutes of Health. Dr. de Vries is funded by R01HL146860 from the National Heart, Lung and Blood Institute (NHLBI). Dr. Wang is supported by the Pioneering Action Grants of the Chinese Academy of Sciences. Molecular data for the Trans Omics in Precision Medicine (TOPMed) program was supported by the NHLBI. Core support including centralized genomic read mapping and genotype calling, along with variant quality metrics and filtering were provided by the TOPMed Informatics Research Center (3R01HL-117626-02S1; contract HHSN268201800002I). Core support including phenotype harmonization, data management, sample-identity QC and general program coordination was provided by the TOPMed Data Coordinating Center (R01HL-120393; U01HL-120393; contract HHSN268201800001I). We gratefully acknowledge the studies and participants who provided biological samples and data for TOPMed. The views expressed in this manuscript are those of the authors and do not necessarily represent the views of the National Heart, Lung, and Blood Institute, the National Institutes of Health, or the U.S. Department of Health and Human Services. We wish to acknowledge the contributions of the consortium working on the development of the NHLBI BioData Catalyst ecosystem. Support for the Genetic Epidemiology Network of Arteriopathy (GENOA) was provided by the National Heart, Lung and Blood Institute (U01 HL054457, U01 HL054464, U01 HL054481, R01 HL119443, and R01 HL087660) of the National Institutes of Health. DNA extraction for “NHLBI TOPMed: Genetic Epidemiology Network of Arteriopathy” (phs001345) was performed at the Mayo Clinic Genotyping Core, and WGS was performed at the DNA Sequencing and Gene Analysis Center at the University of Washington (3R01HL055673-18S1) and the Broad Institute (HHSN268201500014C). We would like to thank the GENOA participants. The Jackson Heart Study (JHS) is supported and conducted in collaboration with Jackson State University (HHSN268201800013I), Tougaloo College (HHSN268201800014I), the Mississippi State Department of Health (HHSN268201800015I) and the University of Mississippi Medical Center (HHSN268201800010I, HHSN268201800011I and HHSN268201800012I) contracts from the NHLBI and the National Institute on Minority Health and Health Disparities (NIMHD). Genome sequencing for “NHLBI TOPMed: The Jackson Heart Study” (phs000964.v1.p1) was performed at the Northwest Genomics Center (HHSN268201100037C). The authors also wish to thank the staffs and participants of the JHS. The MESA projects are conducted and supported by NHLBI in collaboration with MESA investigators. Support for the Multi-Ethnic Study of Atherosclerosis (MESA) projects are conducted and supported by the NHLBI in collaboration with MESA investigators. Support for MESA is provided by contracts 75N92020D00001, HHSN268201500003I, N01-HC-95159, 75N92020D00005, N01-HC-95160, 75N92020D00002, N01-HC-95161, 75N92020D00003, N01-HC-95162, 75N92020D00006, N01-HC-95163, 75N92020D00004, N01-HC-95164, 75N92020D00007, N01-HC-95165, N01-HC-95166, N01-HC-95167, N01-HC-95168, N01-HC-95169, UL1-TR-000040, UL1-TR-001079, UL1-TR-001420, UL1TR001881, DK063491, R01HL105756, and R01HL146860. Genome sequencing for “NHLBI TOPMed: Whole Genome Sequencing and Related Phenotypes in the Multi-Ethnic Study of Atherosclerosis Study (MESA)” (phs001416) was performed at Broad Institute of MIT and Harvard Genomics Platform (3U54HG003067-13S1). The authors thank the other investigators, the staff, and the participants of the MESA study for their valuable contributions. A full list of participating MESA investigators and institutes can be found at http://www.mesa-nhlbi.org. The Women’s Health Initiative (WHI) program is funded by the National Heart, Lung, and Blood Institute, National Institutes of Health, U.S. Department of Health and Human Services through contracts 75N92021D00001, 75N92021D00002, 75N92021D00003, 75N92021D00004, 75N92021D00005. Genome sequencing for “NHLBI TOPMed: Whole Genome Sequencing and Related Phenotypes in the Women’s Health Initiative Study (WHI)” (phs001237) was performed at Broad Institute of MIT and Harvard Genomics Platform (HHSN268201500014C). Support for the Diabetes Heart Study (DHS) by R01 HL92301, R01 HL67348, R01 NS058700, R01 AR48797, R01 DK071891, R01 AG058921, the General Clinical Research Center of the Wake Forest University School of Medicine (M01 RR07122, F32 HL085989), the American Diabetes Association, and a pilot grant from the Claude Pepper Older Americans Independence Center of Wake Forest University Health Sciences (P60 AG10484). Genome sequencing for “NHLBI TOPMed: The Diabetes Heart Study” (phs001412) was performed at the Broad Institute of MIT and Harvard Genomic Platform (HHSN268201500014C). The Atherosclerosis Risk in Communities study has been funded in whole or in part with Federal funds from the National Heart, Lung, and Blood Institute, National Institutes of Health, Department of Health and Human Services, under Contract nos. (75N92022D00001, 75N92022D00002, 75N92022D00003, 75N92022D00004, 75N92022D00005). The authors thank the staff and participants of the ARIC study for their important contributions. Whole genome sequencing (WGS) for the Trans-Omics in Precision Medicine (TOPMed) program was supported by the National Heart, Lung and Blood Institute (NHLBI). WGS for “NHLBI TOPMed: Atherosclerosis Risk in Communities (ARIC)” (phs001211) was performed at the Baylor College of Medicine Human Genome Sequencing Center (HHSN268201500015C and 3U54HG003273-12S2) and the Broad Institute for MIT and Harvard (3R01HL092577-06S1). Centralized read mapping and genotype calling, along with variant quality metrics and filtering were provided by the TOPMed Informatics Research Center (3R01HL-117626-02S1). Phenotype harmonization, data management, sample-identity QC, and general study coordination, were provided by the TOPMed Data Coordinating Center (3R01HL-120393-02S1). We gratefully acknowledge the studies and participants who provided biological samples and data for TOPMed. The Genome Sequencing Program (GSP) was funded by the National Human Genome Research Institute (NHGRI), the National Heart, Lung, and Blood Institute (NHLBI), and the National Eye Institute (NEI). The GSP Coordinating Center (U24 HG008956) contributed to cross program scientific initiatives and provided logistical and general study coordination. The Centers for Common Disease Genomics (CCDG) program was supported by NHGRI and NHLBI, and whole genome sequencing was performed at the Baylor College of Medicine Human Genome Sequencing Center (UM1 HG008898). The COPDGene study (NCT00608764) is supported by grants from the NHLBI (U01HL089897 and U01HL089856), by NIH contract 75N92023D00011, and by the COPD Foundation through contributions made to an Industry Advisory Committee that has included AstraZeneca, Bayer Pharmaceuticals, Boehringer-Ingelheim, Genentech, GlaxoSmithKline, Novartis, Pfizer and Sunovion. A full listing of COPDGene investigators can be found at: http://www.copdgene.org/directory. Genome sequencing for "NHLBI TOPMed: Genetic Epidemiology of COPD Study" (phs000951) was performed at Northwest Genomics Center and Broad Genomics (3R01HL089856-08S1, HHSN268201500014C, HHSN268201500014C). This Cardiovascular Health Study research was supported by NHLBI contracts HHSN268201200036C, HHSN268200800007C, HHSN268201800001C, N01HC55222, N01HC85079, N01HC85080, N01HC85081, N01HC85082, N01HC85083, N01HC85086, 75N92021D00006; and NHLBI grants U01HL080295, R01HL087652, R01HL105756, R01HL103612, R01HL120393, and U01HL130114 with additional contribution from the National Institute of Neurological Disorders and Stroke (NINDS). Additional support was provided through R01AG023629 from the National Institute on Aging (NIA). Genome sequencing for "NHLBI TOPMed: Cardiovascular Health Study" (phs001368.v2.p1) was performed at the Baylor College of Medicine Human Genome Sequencing Center (3U54HG003273-12S2, HHSN268201500015C, HHSN268201600033I). A full list of principal CHS investigators and institutions can be found at CHS-NHLBI.org. The TOPMed component of the Amish Research Program was supported by NIH grants R01 HL121007, U01 HL072515, and R01 AG18728. Genome sequencing for NHLBI TOPMed: Amish (phs000956) was performed at the Broad Institute of MIT and Harvard (3R01HL121007-01S1). The Framingham Heart Study (FHS) acknowledges the support of contracts NO1-HC-25195, HHSN268201500001I and 75N92019D00031 from the National Heart, Lung and Blood Institute and grant supplement R01 HL092577-06S1 for this research. Genome sequencing for “NHLBI TOPMed: Whole Genome Sequencing and Related Phenotypes in the Framingham Heart Study (FHS)” (phs000974) was performed at Broad Institute of MIT and Harvard Genomics Platform (3U54HG003067-12S2). We also acknowledge the dedication of the FHS study participants without whom this research would not be possible. Dr. Vasan is supported in part by the Evans Medical Foundation and the Jay and Louis Coffman Endowment from the Department of Medicine, Boston University School of Medicine. GeneSTAR was supported by the National Institutes of Health/National Heart, Lung, and Blood Institute (U01 HL72518, HL087698, HL112064, HL49762, HL59684, HL58625, HL071025), by the National Institutes of Health/ National Institute of Nursing Research (NR0224103, NR008153), and by a grant from the National Institutes of Health/National Center for Research Resources (M01-RR000052) to the Johns Hopkins General Clinical Research Center. Genome sequencing for NHLBI TOPMed: GeneSTAR (Genetic Study of Atherosclerosis Risk)(phs001218) was performed at the Broad Institute of MIT and Harvard (HHSN268201500014C), at PsomaGen (formerly Macrogen, HHSN268201500014C), and at Illumina (HL112064). We gratefully acknowledge the studies and participants who provided biological samples and data for UK Biobank.

