## supplementary materials for "An Integrated Germline and Somatic Genomic Model Improves Risk Prediction for Coronary Artery Disease"

Pramod Anugu[1], Paul Auer[2], Lucas Barwick[3], Diane Becker[4], Lewis Becker[5], Joshua Bis[6], Cara Carty[7], Peter Castaldi[8], Mark Chaffin[9], Yi-Cheng Chang[10], Seung Hoan Choi[9], Lee-Ming Chuang[11], Ren-Hua Chung[12], Carolyn Crandall[13], Sean David[14], Lisa de las Fuentes[15], Ranjan Deka[16], Dawn DeMeo[17], Qing Duan[18], Charles Eaton[19], Lynette Ekunwe[1], Adel El Boueiz[20], Nora Franceschini[21], Shanshan Gao[22], Yan Gao[1], Margery Gass[23], Auyon Ghosh[17], Daniel Grine[22], Michael Hall[24], Craig Hersh[25], Brian Hobbs[17], Chao (Agnes) Hsiung[26], Yi-Jen Hung[27], Haley Huston[28], Chii Min Hwu[29], Rebecca Jackson[30], Jill Johnsen[31], Christoph Lange[32], Ethan Lange[22], Meryl LeBoff[17], Wen-Jane Lee[29], Yun Li[18], Simin Liu[33], Yu Liu[34], JoAnn Manson[17], Lisa Martin[35], Susan Mathai[22], Hao Mei[1], Rakhi Naik[5], Take Naseri[36], Bonnie Neltner[22], Heather Ochs-Balcom[37], David T. Paik[38], Cora Parker[39], Marco Perez[40], Ulrike Peters[41], Lawrence S Phillips[42], Julia Powers Becker[43], Bruce Psaty[44], Muagututi'a Sefuiva Reupena[45], Carolina Roselli[9], Pamela Russell[22], Ester Cerdeira Sabino[46], Kevin Sandow[47], Karen Schwander[48], Frank Sciurba[49], Brian Silver[50], Sylvia Smoller[51], Beverly Snively[52], Garrett Storm[53], Yun Ju Sung[48], Hua Tang[54], Margaret Taub[5], Lesley Tinker[55], David Tirschwell[44], Hemant Tiwari[56], Dhananjay Vaidya[5], Tarik Walker[22], Robert Wallace[57], Avram Walts[22], Lu-Chen Weng[58], Ivana Yang[22], Snow Xueyan Zhao[59]

[1]University of Mississippi, Jackson, Mississippi, United States of America, [2]Medical College of Wisconsin, Milwaukee, Wisconsin, United States of America, [3]LTRC, The Emmes Corporation, Rockville, Maryland, United States of America, [4]Medicine, Johns Hopkins University, Baltimore, Maryland, United States of America, [5]Johns Hopkins University, Baltimore, Maryland, United States of America, [6]Cardiovascular Health Research Unit, Department of Medicine, University of Washington, Seattle, Washington, United States of America, [7]Washington State University, Pullman, Washington, United States of America, [8]Medicine, Brigham & Women's Hospital, Boston, Massachusetts, United States of America, [9]Department of Biostatistics, Boston University, Boston, Massachusetts, United States of America, [10]National Taiwan University, Taipei, Taiwan (Province of China), [11]National Taiwan University Hospital, National Taiwan University, Taipei, Taiwan (Province of China), [12]National Health Research Institute Taiwan, Miaoli County, Taiwan (Province of China), [13]University of California, Los Angeles, Los Angeles, California, United States of America, [14]University of Chicago, Chicago, Illinois, United States of America, [15]Department of Medicine, Cardiovascular Division, Washington University in St Louis, St. Louis, Missouri, United States of America, [16]University of Cincinnati, Cincinnati, Ohio, United States of America, [17]Brigham & Women's Hospital, Boston, Massachusetts, United States of America, [18]University of North Carolina, Chapel Hill, North Carolina, United States of America, [19]Brown University, Providence, Rhode Island, United States of America, [20]Channing Division of Network Medicine, Harvard University, Cambridge, Massachusetts, United States of America, [21]Epidemiology, University of North Carolina, Chapel Hill, North Carolina, United States of America, [22]University of Colorado at Denver, Denver, Colorado, United States of America, [23]Fred Hutchinson Cancer Research Center, Seattle, Washington, United States of America, [24]Cardiology, University of Mississippi, Jackson, Mississippi, United States of America, [25]Channing Division of Network Medicine, Brigham & Women's Hospital, Boston, Massachusetts, United States of America, [26]Institute of Population Health Sciences, NHRI, National Health Research Institute Taiwan, Miaoli County, Taiwan (Province of China), [27]Tri-Service General Hospital National Defense Medical Center, Taiwan (Province of China), [28]Blood Works Northwest, Seattle, Washington, United States of America, [29]Taichung Veterans General Hospital Taiwan, Taichung City, Taiwan (Province of China), [30]Internal Medicine, DIvision of Endocrinology, Diabetes and Metabolism, Oklahoma State University Medical Center, Columbus, Ohio, United States of America, [31]Medicine, University of Washington, Seattle, Washington, United States of America, [32]Biostats, Harvard School of Public Health, Boston, Massachusetts, United States of America, [33]Epidemiology and Medicine, Brown University, Providence, Rhode Island, United States of America, [34]Cardiovascular Institute, Stanford University, Stanford, California, United States of America, [35]cardiology, George Washington University, Washington, District of Columbia, United States of America, [36]Ministry of Health, Government of Samoa, Apia, Samoa, [37]University at Buffalo, Buffalo, New York, United States of America, [38]Stanford Cardiovascular Institute, Stanford University, Stanford, California, United States of America, [39]Biostatistics and Epidemiology Division, RTI International, Research Triangle Park, North Carolina, United States of America, [40]Stanford University, Stanford, California, United States of America, [41]Fred Hutch and UW, Fred Hutchinson Cancer Research Center, Seattle, Washington, United States of America, [42]Emory University, Atlanta, Georgia, United States of America, [43]Medicine, University of Colorado at Denver, Denver, Colorado, United States of America, [44]University of Washington, Seattle,

Washington, United States of America, [45]Lutia I Puava Ae Mapu I Fagalele, Apia, Samoa, [46]Faculdade de Medicina, Universidade de Sao Paulo, Sao Paulo, Brazil, [47]TGPS, Lundquist Institute, Torrance, California, United States of America, [48]Washington University in St Louis, St Louis, Missouri, United States of America, [49]University of Pittsburgh, Pittsburgh, Pennsylvania, United States of America, [50]UMass Memorial Medical Center, Worcester, Massachusetts, United States of America, [51]Albert Einstein College of Medicine, New York, New York, United States of America, [52]Biostatistical Sciences, Wake Forest Baptist Health, Winston-Salem, North Carolina, United States of America, [53]Genomic Cardiology, University of Colorado at Denver, Aurora, Colorado, United States of America, [54]Genetics, Stanford University, Stanford, California, United States of America, [55]Cancer Prevention Division of Public Health Sciences, Fred Hutchinson Cancer Research Center, Seattle, Washington, United States of America, [56]Biostatistics, University of Alabama, Birmingham, Alabama, United States of America, [57]University of Iowa, Iowa City, Iowa, United States of America, [58]Massachusetts General Hospital, Boston, Massachusetts, United States of America, [59]National Jewish Health, Denver, Colorado, United States of America

**Supplementary Table 1** Participant characteristics for UK Biobank and TOPMed

| | **UK Biobank** | **TOPMED** |
|---|---|---|
| **n** | 391536 | 34177 |
| **Age, mean (SD)** | 56.5 (8.1) | 62.6 (10.6) |
| **Male (%)** | 181014 (46.2) | 11624 (34.0) |
| **Follow-up duration, years, median (IQR)** | 12.3 (1.6) | 10.5 (8.6) |
| **Smoking, n (%)** | | |
| Missing | 1586 (0.4) | - |
| Never | 213818 (54.7) | 12023 (35.2) |
| Previous | 135049 (34.5) | 13775 (40.3) |
| Current | 40651 (10.4) | 8380 (24.5) |
| **CAD, n (%)** | 42602 (10.9) | 6421 (18.8) |
| Incident CAD (%) | 28346 (7.2) | 4326 (12.7) |
| **Race/ethnicity, n (%)** | | |
| Asian | 9106 (2.3) | 737 (2.2) |
| Black | 6237 (1.6) | 8858 (25.9) |
| Other | 7897 (2.0) | 1250 (3.7) |
| White | 368296 (94.1) | 23333 (68.3) |
| **BMI, kg/m2, mean (SD)** | 27.43 (4.77) | 28.5 (6.00) |
| **Measured DBP, mmHg, mean (SD)** | 82.22 (10.68) | - |
| **Measured SBP, mmHg, mean (SD)** | 139.67 (19.59) | - |
| **Measured TC, mmol/L, mean (SD)** | 5.69 (1.14) | 5.52 (1.10) |
| **Measured TG, mmol/L, mean (SD)** | 1.75 (1.03) | 1.53 (0.99) |
| **Measured HDL, mmol/L, mean (SD)** | 1.45 (0.38) | 1.37 (0.40) |
| **Measured LDL, mmol/L, mean (SD)** | 3.56 (0.87) | 3.45 (0.98) |

IQR: interquartile range

**Supplementary Table 2** Characteristics of FH carriers and noncarriers in UK Biobank

| | **FH non-carries** | **FH carriers** |
|---|---|---|
| **n** | 390433 | 1103 |
| **Male, n (%)** | 180542(46.2) | 472(42.8) |
| **Smoking, n (%)** | | |
| Missing | 1579(0.4) | 7(0.6) |
| Never | 213224(54.7) | 594(54.0) |
| Previous | 134653(34.5) | 396(36.0) |
| Current | 40547(10.4) | 104(9.4) |
| **CAD, n (%)** | 42391(10.9) | 211(19.1) |
| Incident CAD | 28230(7.2) | 116(10.5) |
| **Race/ethnicity, n (%)** | | |
| Asian | 9083(2.3) | 23(2.1) |
| Black | 6224(1.6) | 13(1.2) |
| Other | 7883(2.0) | 14(1.3) |
| White | 367243(94.1) | 1053(95.5) |
| **Age, mean (SD)** | 56.47(8.05) | 56.56(8.03) |
| **BMI, kg/m2, mean (SD)** | 27.43(4.77) | 27.51(4.93) |
| **Estimated untreated DBP, mmHg, mean (SD)** | 83.36(11.43) | 81.96(11.79) |
| **Estimated untreated SBP, mmHg, mean (SD)** | 141.38(20.82) | 140.37(20.85) |
| **Estimated untreated TC, mmol/L, mean (SD)** | 5.89(1.09) | 7.19(1.52) |
| **Estimated untreated TG, mmol/L, mean (SD)** | 1.81(1.09) | 1.73(1.10) |
| **Estimated untreated HDL, mmol/L, mean (SD)** | 1.45(0.38) | 1.41(0.35) |
| **Estimated untreated LDL, mmol/L, mean (SD)** | 3.76(0.85) | 5.05(1.26) |

Untreated blood pressure was estimated by adjusting the raw value for anti-hypertensive medication intake by adding 15 mmHg to the systolic blood pressure and 10 mmHg to the diastolic blood pressure; lipids were adjusted by the formula from Supplementary Table 19.

**Supplementary Table 3** Characteristics of FH carriers and noncarriers in TOPMed

| | **FH non-carrier** | **FH carrier** |
|---|---|---|
| **n** | 34093 | 85 |
| **Age, mean (SD)** | 62.6 (10.6) | 60.8 (12.1) |
| **Male (%)** | 11601 (34.0) | 23 (27.1) |
| **Smoking, n (%)** | | |
| Never | 11993 (35.2) | 30 (35.3) |
| Previous | 13737 (40.3) | 38 (44.7) |
| Current | 8363 (24.5) | 17 (20.0) |
| **CAD, n (%)** | 6392 (18.7) | 29 (34.1) |
| Incident CAD (%) | 4312 (12.6) | 14 (16.5) |
| **Race/ethnicity, n (%)** | | |
| Asian | 735 (2.2) | 2 (2.4) |
| Black | 8839 (25.9) | 19 (22.4) |
| Other | 1248 (3.7) | 2 (2.4) |
| White | 23271 (68.3) | 62 (72.9) |
| **BMI, kg/m2, mean (SD)** | 28.5 (6.00) | 28.9 (6.86) |
| **Measured TC, mmol/L, mean (SD)** | 5.52 (1.09) | 6.73 (2.07) |
| **Measured TG, mmol/L, mean (SD)** | 1.54 (0.99) | 1.48 (0.98) |
| **Measured HDL, mmol/L, mean (SD)** | 1.37 (0.40) | 1.28 (0.41) |
| **Measured LDL, mmol/L, mean (SD)** | 3.45 (0.98) | 4.72 (2.08) |

**Supplementary Table 4** Characteristics of CHIP carriers and noncarriers in UK Biobank

| | **CHIP Non-Carries** | **CHIP Carriers** |
|---|---|---|
| **n** | 382120 | 9416 |
| **Male, n (%)** | 175952(46.0) | 5062(53.8) |
| **Smoking, n (%)** | | |
| Missing | 1526(0.4) | 60(0.6) |
| Never | 209409(54.9) | 4409(46.9) |
| Previous | 131291(34.4) | 3758(39.9) |
| Current | 39469(10.3) | 1182(12.6) |
| **CAD, n (%)** | 41021(10.7) | 1581(16.8) |
| Incident CAD, n (%) | 27343(7.2) | 1003(10.7) |
| **Race/ethnicity, n (%)** | | |
| Asian | 8939(2.3) | 167(1.8) |
| Black | 6147(1.6) | 90(1.0) |
| Other | 7746(2.0) | 151(1.6) |
| White | 359288(94.0) | 9008(95.7) |
| **Age, mean (SD)** | 56.37(8.04) | 60.61(6.91) |
| **BMI, kg/m2, mean (SD)** | 27.42(4.78) | 27.78(4.68) |
| **Estimated untreated DBP, mmHg, mean (SD)** | 83.34(11.42) | 84.19(11.51) |
| **Estimated untreated SBP, mmHg, mean (SD)** | 141.26(20.80) | 145.83(21.13) |
| **Estimated untreated TC, mmol/L, mean (SD)** | 5.90(1.09) | 5.86(1.10) |
| **Estimated untreated TG, mmol/L, mean (SD)** | 1.81(1.09) | 1.90(1.11) |
| **Estimated untreated HDL, mmol/L, mean (SD)** | 1.45(0.38) | 1.40(0.38) |
| **Estimated untreated LDL, mmol/L, mean (SD)** | 3.76(0.86) | 3.77(0.85) |

Untreated blood pressure was estimated by adjusting the raw value for anti-hypertensive medication intake by adding 15 mmHg to the systolic blood pressure and 10 mmHg to the diastolic blood pressure; lipids were adjusted by the formula from Supplementary Table 19.

**Supplementary Table 5** Characteristics of CHIP carriers and noncarriers in TOPMed

| | **CHIP non-carrier** | **CHIP carrier** |
|---|---|---|
| **n** | 33258 | 920 |
| **Age, mean (SD)** | 62.4 (10.5) | 70.1 (8.53) |
| **Male, n (%)** | 11294 (34.0) | 330 (35.9) |
| **Smoking, n (%)** | | |
| Never | 11696 (35.2) | 327 (35.5) |
| Previous | 13329 (40.1) | 446 (48.5) |
| Current | 8233 (24.8) | 147 (16.0) |
| **Incident CAD, n (%)** | 4156 (12.5) | 170 (18.5) |
| **Race/ethnicity, n (%)** | | |
| Asian | 728 (2.2) | 9 (1.0) |
| Black | 8720 (26.2) | 138 (15.0) |
| Other | 1225 (3.7) | 25 (2.7) |
| White | 22585 (67.9) | 748 (81.3) |
| **BMI, kg/m2, mean (SD)** | 28.5 (6.00) | 27.9 (5.83) |
| **Measured TC, mmol/L, mean (SD)** | 5.52 (1.10) | 5.60 (1.14) |
| **Measured TG, mmol/L, mean (SD)** | 1.53 (0.99) | 1.63 (0.97) |
| **Measured HDL, mmol/L, mean (SD)** | 1.37 (0.40) | 1.37 (0.40) |
| **Measured LDL, mmol/L, mean (SD)** | 3.45 (0.98) | 3.48 (0.98) |

**Supplementary Table 6** Association of germline and somatic genomic drivers with prevalent CAD in the UK Biobank

| **Drivers** | **OR** | **OR %95CI** | **P-Value** |
|---|---|---|---|
| GermRisk | 2.158 | (2.119,2.199) | <0.0001 |
| **PRS** | 2.149 | (2.110,2.190) | <0.0001 |
| **MetPRS** | 1.273 | (1.251,1.295) | <0.0001 |
| **ProPRS** | 1.191 | (1.170,1.212) | <0.0001 |
| **FH** | 3.079 | (2.461,3.853) | <0.0001 |
| *LDLR* | 3.414 | (2.630,4.432) | <0.0001 |
| *APOB* | 2.157 | (1.340,3.473) | 0.0016 |
| *PCSK9* | 4.499 | (1.475,13.719) | 0.0082 |
| hGermRisk[#] | 3.595 | (3.469,3.726) | <0.0001 |
| **hPRS**[#] | 3.521 | (3.398,3.650) | <0.0001 |
| **hMetPRS**[#] | 1.546 | (1.487,1.608) | <0.0001 |
| **hProPRS**[#] | 1.333 | (1.280,1.388) | <0.0001 |

Modeling each factor independently by a logistic regression model, corrected for covariates sex, age and top 10 PCs, prevalent cases were used. [#]PRS, MetPRS, and ProPRS were binarized, with individuals in the top 20% treated as carriers of hPRS, hMetPRS, hProPRS, respectively. PRS: CAD PRS; ProPRS: a proteome PRS trained in this study; MetPRS a metabolome PRS trained in this study; FH: familial hypercholesterolemia. OR, odds ratio per standard deviation for each continuous measure, and odds ratio per carrier status for FH and CHIP, respectively.

**Supplementary Table 7** Association of germline and somatic genomic drivers with prevalent CAD in TOPMed

| Drivers | **OR** | **OR %95CI** | **P-Value** |
|---|---|---|---|
| GermRisk | 1.608 | (1.561, 1.657) | <0.0001 |
| **PRS** | 1.603 | (1.555, 1.651) | <0.0001 |
| **MetPRS** | 1.143 | (1.111, 1.176) | <0.0001 |
| **ProPRS** | 1.068 | (1.039, 1.099) | <0.0001 |
| **FH** | 2.656 | (1.852, 4.289) | <0.0001 |
| hGermRisk[#] | 2.230 | (2.088, 2.382) | <0.0001 |
| **hPRS**[#] | 2.222 | (2.080, 2,373) | <0.0001 |
| **hMetPRS**[#] | 1.263 | (1.178, 1.354) | <0.0001 |
| **hProPRS**[#] | 1.132 | (1.056, 1.214) | <0.0001 |

Modeling each factor independently by a logistic regression model, corrected for covariates sex, age, and top 10 PCs, prevalent cases were used. [#]PRS, MetPRS, and ProPRS were binarized, with individuals in the top 20% treated as carriers of hPRS, hMetPRS, hProPRS, respectively. PRS: CAD PRS; ProPRS: a proteome PRS trained in this study; MetPRS a metabolome PRS trained in this study; FH: familial hypercholesterolemia. OR, odds ratio per standard deviation for each continuous measure, and odds ratio per carrier status for FH and CHIP, respectively.

**Supplementary Table 8** Association of germline and somatic genomic drivers with incident CAD in the UK Biobank

| Drivers | **HR** | **HR %95CI** | **P-Value** |
|---|---|---|---|
| IGMRisk | 1.576 | (1.558,1.595) | <0.0001 |
| GermRisk | 1.574 | (1.555,1.593) | <0.0001 |
| **PRS** | 1.564 | (1.546,1.583) | <0.0001 |
| **MetPRS** | 1.180 | (1.166,1.194) | <0.0001 |
| **ProPRS** | 1.153 | (1.140,1.167) | <0.0001 |
| **FH** | 1.689 | (1.407,2.027) | <0.0001 |
| *LDLR* | 1.776 | (1.432,2.203) | <0.0001 |
| *APOB* | 1.413 | (0.976,2.047) | 0.0671 |
| *PCSK9* | 2.282 | (0.949,5.483) | 0.0653 |
| SomaRisk | 1.052 | (1.042,1.063) | <0.0001 |
| **LTL** | 0.936 | (0.925,0.947) | <0.0001 |
| **CHIP** | 1.127 | (1.058,1.201) | 0.0002 |
| hIGMRisk# | 2.167 | (2.114,2.221) | <0.0001 |
| hGermRisk# | 2.147 | (2.095,2.201) | <0.0001 |
| **hPRS**# | 2.126 | (2.073,2.179) | <0.0001 |
| **hMetPRS**# | 1.342 | (1.306,1.379) | <0.0001 |
| **hProPRS**# | 1.281 | (1.246,1.316) | <0.0001 |
| **FH** | 1.689 | (1.407,2.027) | <0.0001 |
| hSomaRisk# | 1.142 | (1.110,1.174) | <0.0001 |
| **sLTL**$ | 1.143 | (1.111,1.175) | <0.0001 |
| **CHIP** | 1.127 | (1.058,1.201) | 0.0002 |

Modeling each factor independently, corrected for covariates sex, age and top 10 PCs. # indicates top 20% of each variable; $ indicates bottom 20%. PRS, MetPRS, ProPRS, and LTL were binarized, with individuals in the top 20% treated as carriers (hPRS, hMetPRS, hProPRS) or bottom 20% treated as carriers (sLTL). HR, hazard ratio per standard deviation for each continuous measure, and hazard ratio per carrier status for FH and CHIP, respectively.

**Supplementary Table 9** Association of germline and somatic genomic drivers with incident CAD in TOPMed

| Drivers | **HR** | **HR %95CI** | **P-Value** |
|---|---|---|---|
| IGMRisk | 1.464 | (1.403, 1.528) | <0.0001 |
| GermRisk | 1.458 | (1.404, 1.515) | <0.0001 |
| **PRS** | 1.468 | (1.411, 1.528) | <0.0001 |
| **MetPRS** | 1.103 | (1.088, 1.119) | <0.0001 |
| **ProPRS** | 1.048 | (1.029, 1.067) | <0.0001 |
| **FH** | 1.790 | (1.322, 2.423) | <0.0001 |
| SomaRisk | 1.053 | (1.017, 1.090) | 0.0033 |
| **LTL** | 0.939 | (0.898, 0.982) | 0.0053 |
| **CHIP** | 1.159 | (0.998, 1.345) | 0.0534 |
| hIGMRisk$^{\#}$ | 1.924 | (1.790, 2.068) | <0.001 |
| hGermRisk$^{\#}$ | 1.937 | (1.833, 2.047) | <0.0001 |
| **hPRS**$^{\#}$ | 1.914 | (1.815, 2.018) | <0.0001 |
| **hMetPRS**$^{\#}$ | 1.180 | (1.157, 1.204) | <0.0001 |
| **hProPRS**$^{\#}$ | 1.092 | (1.062, 1.123) | <0.0001 |
| **FH** | 1.790 | (1.322, 2.423) | <0.0001 |
| hSomaRisk$^{\#}$ | 1.207 | (1.088, 1.339) | <0.0001 |
| **sLTL**$^{\$}$ | 1.164 | (1.052, 1.289) | 0.0033 |
| **CHIP** | 1.159 | (0.998, 1.345) | 0.0534 |

Modeling each factor independently, corrected for covariates sex, age and top 10 PCs. $^{\#}$ indicates top 20% of each variable; $^{\$}$ indicates bottom 20%. PRS, MetPRS, ProPRS, and LTL were binarized, with individuals in the top 20% treated as carriers (hPRS, hMetPRS, hProPRS) or bottom 20% treated as carriers (sLTL). HR, hazard ratio per standard deviation for each continuous measure, and hazard ratio per carrier status for FH and CHIP, respectively.

**Supplementary Table 10** Composition of genomic drivers in different IGM risk strata

| **UK Biobank** | | |
|---|---|---|
| | **Count (%)** | **10-Year risk of CAD (%)(95% CI)** |
| **Low risk IGM group** | 75454 (20%) | 2.22 (2.15, 2.28) |
| **Intermediate risk IGM group** | 226368 (60%) | 4.12 (4.06, 4.19) |
| **High risk IGM group** | 75454 (20%) | 7.60 (7.45, 7.76) |
| ● Non-high germline and somatic risk* | 624 (0.8%)† | 5.93 (5.38,6.47) |
| ● Non-high germline and high somatic risk* | 4527 (6.0%)† | 6.57 (6.04,7.10) |
| ● High germline and non-high somatic risk* | 54785 (72.6%)† | 7.96 (7.74,8.18) |
| ● High germline and somatic risk* | 15520 (20.6%)† | 8.81 (8.43,9.19) |
| **TOPMed** | | |
| | Count (%) | 10-Year risk of CAD (%)(95% CI) |
| **Low risk IGM group** | 5504 (20%) | 6.70 (6.23,7.18) |
| **Intermediate risk IGM group** | 16512 (60%) | 11.20 (10.72,11.67) |
| **High risk IGM group** | 5504 (20%) | 18.40 (17.41,19.37) |
| ● Non-high germline and somatic risk* | 54 (1.0%)† | 13.14 (9.55,16.58) |
| ● Non-high germline and high somatic risk* | 313 (5.7%)† | 15.01 (11.38,18.5) |
| ● High germline and non-high somatic risk* | 4016 (73.0%)† | 19.36 (17.9,20.8) |
| ● High germline and somatic risk* | 1121 (20.4%)† | 22.01 (19.36,24.57) |

The ten-year risk of CAD was estimated for each IGM risk group in the UK Biobank dataset. Categories were defined as low risk (bottom 20%), intermediate risk (middle 60%), and high risk group (top 20%). In the high risk group, the genetic profile was further partitioned by the status of carrying a high germline risk or high somatic risk. The high germline risk was defined as the top 20% with a composite risk estimated from four germline genetic risk drivers (FH, PRS, MetPRS, and ProPRS). The high somatic risk was defined as the top 20% with a composite risk estimated from two somatic risk drivers (CHIP and LTL). PRS, polygenic risk score (PRS) for CAD; MetPRS, metabolome PRS; ProPRS, proteome PRS; LTL, leukocyte telomere length; FH, familial hypercholesterolemia variants; CHIP, clonal hematopoiesis of indeterminate potential. *Combinations in high risk group; †Percentage among high risk group.

**Supplementary Table 11** Number of genomic drivers in high and low risk groups identified by IGM

| **Genetic combinations among high risk groups (top 20%) identified by IGM** | **Count (%) in UK Biobank** | **Count (%) in TOPMed** |
|---|---|---|
| High risk for 6 genetic drivers | 0 (0) | 0 (0) |
| High risk for 5 genetic drivers | 45 (0.06) | 6 (0.11) |
| High risk for 4 genetic drivers | 2026 (2.69) | 128 (2.33) |
| High risk for 3 genetic drivers | 13707 (18.17) | 957 (17.39) |
| High risk for 2 genetic drivers | 32412 (42.95) | 2340 (42.51) |
| High risk for 1 genetic driver | 27015 (35.80) | 2057 (37.37) |
| High risk for 0 genetic drivers | 251 (0.33) | 16 (0.29) |
| **Genetic combinations among low risk group (bottom 20%) identified by IGM** | Count (%) in UK Biobank | Count (%) in TOPMed |
| High risk for 6 genetic drivers | 0 (0) | 0 (0) |
| High risk for 5 genetic drivers | 0 (0) | 0 (0) |
| High risk for 4 genetic drivers | 0 (0) | 0 (0) |
| High risk for 3 genetic drivers | 134 (0.18) | 9 (0.16) |
| High risk for 2 genetic drivers | 2409 (3.19) | 184 (3.34) |
| High risk for 1 genetic driver | 19569 (25.93) | 1488 (27.03) |
| High risk for 0 genetic drivers | 53344 (70.70) | 3823 (69.46) |

The participants were defined to have genetic drivers for PRS, ProPRS, and MetPRS when they were among the 20%, and for LTL when they were among the bottom 20% risk. Six drivers of FH, PRS, MetPRS, ProPRS, CHIP, and LTL were counted. PRS, polygenic risk score (PRS) for CAD; MetPRS, metabolome PRS; ProPRS, proteome PRS; LTL, leukocyte telomere length; FH, familial hypercholesterolemia variants; CHIP, clonal hematopoiesis of indeterminate potential.

**Supplementary Table 12** Stratification table of PCE vs PCE+IGM in UK Biobank

| PCE | PCE+IGM | | | | Total |
|---|---|---|---|---|---|
| | **Low (<5%)** | **Borderline (5% to 7.5%)** | **Intermediate (7.5% to 20%)** | **High (>20%)** | |
| **Low (<5%)** | | | | | |
| Sample, n | 196746 | 14923 | 3076 | 0 | 214745 |
| Event, n | 3661 | 928 | 285 | 0 | 4874 |
| Nonevent, n | 193085 | 13995 | 2791 | 0 | 209871 |
| Event proportion | 1.9% | 6.2% | 9.3% | NA | 2.3% |
| **Borderline (5% to 7.5%)** | | | | | |
| Sample, n | 23313 | 24127 | 15456 | 34 | 62930 |
| Event, n | 872 | 1504 | 1561 | 12 | 3949 |
| Nonevent, n | 22441 | 22623 | 13895 | 22 | 58981 |
| Event proportion | 3.7% | 6.2% | 10.1% | 35.3% | 6.3% |
| **Intermediate (7.5% to 20%)** | | | | | |
| Sample, n | 5500 | 18897 | 65165 | 6221 | 95783 |
| Event, n | 230 | 1221 | 7983 | 1469 | 10903 |
| Nonevent, n | 5270 | 17676 | 57182 | 4752 | 84880 |
| Event proportion | 4.2% | 6.5% | 12.3% | 23.6% | 11.4% |
| **High (>20%)** | | | | | |
| Sample, n | 0 | 9 | 1464 | 2349 | 3822 |
| Event, n | 0 | 1 | 260 | 637 | 898 |
| Nonevent, n | 0 | 8 | 1204 | 1712 | 2924 |
| Event proportion | NA | 11.1% | 17.8% | 27.1% | 23.5% |
| **Total** | | | | | |
| Sample, n | 225559 | 57956 | 85161 | 8604 | 377280 |
| Event, n | 4763 | 3654 | 10089 | 2118 | 20624 |
| Nonevent, n | 220796 | 54302 | 75072 | 6486 | 356656 |
| Event proportion | 2.1% | 6.3% | 11.8% | 24.6% | 5.5% |

**Supplementary Table 13** C-statistic evaluating the performance of the IGM in UK Biobank and TOPMed

| **UK Biobank*** | |
|---|---|
| **Model** | **C-statistic (95% CI)** |
| Base model[a] | 0.701 (0.698–0.704) |
| Clinical risk model[b] | 0.725 (0.723–0.728) |
| Integrated genomic model[c] | 0.734 (0.731–0.736) |
| Integrated genomic and clinical risk model[d] | 0.750 (0.747–0.753) |
| **TOPMed†** | |
| **Model** | **C-statistic (95% CI)** |
| Base model[a] | 0.674 (0.666–0.683) |
| Integrated genomic model[c] | 0.707 (0.699–0.715) |
| Integrated genomic model (age $\leq$50)[c] | 0.725 (0.683–0.767) |
| Integrated genomic model (age $\leq$45)[c] | 0.805 (0.699–0.913) |

a: Age, sex, top 10 principal components (PC).
b: Pooled cohort equation, natural log transformed.
c: Base model, integrated genomic model (germline risk and somatic risk).
d: Base model, integrated genomic model, and natural log PCE.
*Age stratified analysis was not performed in the UK Biobank data set, considering that all participants were enrolled at ages over 40.
†Clinical risk model was not assessed in TOPMed as covariates necessary for PCE calculation were absent for most participants in the cohort.

**Supplementary Table 14** Enrichment analysis of integrated genomic risk in UK Biobank

| **UK Biobank** | | | | |
|---|---|---|---|---|
| | **Integrated genomic risk category** | | | |
| | **Bottom 5%** | **Bottom 20%** | **Top 20%** | **Top 5%** |
| **No. of samples** | 18864 | 75456 | 75456 | 18864 |
| **No. of Case** | 607 (3.22) | 3003 (3.98) | 9553 (12.66) | 3084 (16.35) |
| **No. of FH carriers** | 4 (0.02) | 32 (0.04) | 609 (0.81) | 299 (1.59) |
| **No. of CHIP carriers** | 88 (0.47) | 593 (0.79) | 3888 (5.15) | 1424 (7.55) |
| **hPRS** | 0 (0) | 0 (0) | 65752 (87.14) | 18694 (99.1) |
| **hMetPRS** | 809 (4.29) | 5744 (7.61) | 27083 (35.89) | 8688 (46.06) |
| **hProPRS** | 957 (5.07) | 6563 (8.7) | 25311 (33.54) | 7751 (41.09) |
| **sLTL** | 2628 (13.93) | 11857 (15.71) | 18646 (24.71) | 5161 (27.36) |
| **TOPMed** | | | | |
| | **Integrated genomic risk category** | | | |
| | **Bottom 5%** | **Bottom 20%** | **Top 20%** | **Top 5%** |
| **No. of samples** | 1376 | 5504 | 5504 | 1376 |
| **No. of Case** | 95 (6.9) | 498 (9.1) | 1200 (21.8) | 373 (27.1) |
| **No. of FH carriers** | 0 (0.0) | 0 (0.0) | 36 (0.0) | 17 (0.0) |
| **No. of CHIP carriers** | 4 (0.3) | 48 (0.9) | 306 (5.6) | 116 (8.4) |
| **hPRS** | 0 (0.0) | 0 (0.0) | 4823 (87.6) | 1367 (99.3) |
| **hMetPRS** | 65 (4.7) | 417 (7.6) | 1881 (34.2) | 598 (43.5) |
| **hProPRS** | 85 (6.2) | 530 (9.6) | 1757 (31.9) | 542 (39.4) |
| **sLTL** | 192 (14.0) | 888 (16.1) | 1347 (24.5) | 368 (26.7) |

Numbers in parentheses represent the percentage of each carrier group relative to the total number of samples within the respective vertical categories (e.g., Top 5%), Some individuals may have more than one risk factor. PRS, MetPRS, ProPRS, and LTL were binarized, with individuals in the top 20% treated as carriers (hPRS, hMetPRS, hProPRS) or bottom 20% treated as carriers (sLTL).

**Supplementary Table 15** Genetic risk score for 124 proteins were retained in the ProPRS lasso models for CAD prediction

| OMICSPRED_ID | Weight | Gene | Protein |
|---|---|---|---|
| OPGS000024 | -0.005258304952 | *MBL2* | Mannose-binding protein C |
| OPGS000031 | 0.008762377743 | *AAGAB* | Alpha- and gamma-adaptin-binding protein p34 |
| OPGS000034 | -0.00337685549 | *SAA1* | Serum amyloid A-1 protein |
| OPGS000044 | 0.0115457839 | *CTRB2* | Chymotrypsinogen B2 |
| OPGS000056 | -0.001085813597 | *MANEA* | Glycoprotein endo-alpha-1,2-mannosidase |
| OPGS000062 | 0.00520278987 | *MSMB* | Beta-microseminoprotein |
| OPGS000092 | -0.003023672673 | *HGFAC* | Hepatocyte growth factor activator |
| OPGS000094 | -0.003061624085 | *PDGFRB* | Platelet-derived growth factor receptor beta |
| OPGS000119 | -0.0149303476 | *IDUA* | Alpha-L-iduronidase |
| OPGS000122 | -0.001539341356 | *PTGFRN* | Prostaglandin F2 receptor negative regulator |
| OPGS000137 | -0.008584003344 | *C4A\|C4B* | Complement C4b |
| OPGS000141 | 0.02844547739 | *C1S* | Complement C1s subcomponent |
| OPGS000154 | 0.01286122983 | *GRN* | Granulins |
| OPGS000170 | -0.009533530839 | *LCT* | Lactase-phlorizin hydrolase |
| OPGS000179 | 0.011586131 | *CTF1* | Cardiotrophin-1 |
| OPGS000210 | -0.001164890135 | *DRGX* | Dorsal root ganglia homeobox protein |
| OPGS000229 | 0.03486879446 | *IL19* | Interleukin-19 |
| OPGS000248 | -0.002705532087 | *CCL23* | Ck-beta-8-1 |
| OPGS000283 | 0.000995697454 | *CAMP* | Cathelicidin antimicrobial peptide |
| OPGS000301 | -0.01072192977 | *HAVCR2* | Hepatitis A virus cellular receptor 2 |
| OPGS000312 | 0.05917550434 | *SELENOS* | Selenoprotein S |
| OPGS000349 | 0.0005420345107 | *GGH* | Gamma-glutamyl hydrolase |
| OPGS000367 | 0.003261882222 | *PDGFRA* | Platelet-derived growth factor receptor alpha |
| OPGS000383 | 0.03586331043 | *FAM3B* | Protein FAM3B |
| OPGS000384 | -0.02104031541 | *ENTPD5* | Ectonucleoside triphosphate diphosphohydrolase 5 |
| OPGS000399 | -0.005678638937 | *GZMB* | Granzyme B |
| OPGS000417 | -0.02180946826 | *B3GNT8* | UDP-GlcNAc:betaGal beta-1,3-N-acetylglucosaminyltransferase 8 |
| OPGS000440 | 0.001161037742 | *IGLL1* | Immunoglobulin lambda-like polypeptide 1 |
| OPGS000491 | -0.01080423524 | *NTN1* | Netrin-1 |
| OPGS000533 | -0.01743560713 | *SPOCK3* | Testican-3 |
| OPGS000577 | 0.02383673934 | *PLA2G7* | Platelet-activating factor acetylhydrolase |
| OPGS000584 | 0.006567180714 | *CCL21* | C-C motif chemokine 21 |
| OPGS000626 | -0.01232384609 | *CD300A* | CMRF35-like molecule 8 |
| OPGS000642 | -0.007464831067 | *SERPIND1* | Heparin cofactor 2 |
| OPGS000653 | 0.02641435999 | *NPW* | Neuropeptide W |
| OPGS000713 | -0.02155941229 | *TIMP4* | Metalloproteinase inhibitor 4 |
| OPGS000719 | 0.01734814721 | *PTHLH* | Parathyroid hormone-related protein |
| OPGS000758 | 0.004737035052 | *STAB1* | Stabilin-1 |
| OPGS000782 | -0.0159165426 | *B4GALT2* | Beta-1,4-galactosyltransferase 2 |
| OPGS000799 | -0.002696573104 | *FYN* | Tyrosine-protein kinase Fyn |
| OPGS000807 | -0.0063023105 | *CCL22* | C-C motif chemokine 22 |
| OPGS000836 | -0.001208500117 | *XCL1* | Lymphotactin |
| OPGS000883 | 0.09347695832 | *PLA2G12B* | Group XIIB secretory phospholipase A2-like protein |

| | | | |
|---|---|---|---|
| OPGS000935 | -0.02702587123 | *VTN* | Vitronectin |
| OPGS001003 | -0.02077909303 | *ALPL* | Alkaline phosphatase, tissue-nonspecific isozyme |
| OPGS001007 | -0.05052743536 | *NRP1* | Neuropilin-1 |
| OPGS001014 | -0.07414068979 | *THBS4* | Thrombospondin-4 |
| OPGS001046 | -0.009338972569 | *NME2* | Nucleoside diphosphate kinase B |
| OPGS001096 | 0.03658497129 | *FAM151A* | Protein FAM151A |
| OPGS001098 | 0.002115173612 | *DYNLRB1* | Dynein light chain roadblock-type 1 |
| OPGS001120 | 0.00324987376 | *IGHG1\|IGHG2\|IGHG3\|IGHG4\|IGK\|IGL* | Immunoglobulin G |
| OPGS001132 | -0.004304528278 | *NPPB* | N-terminal pro-BNP |
| OPGS001144 | 0.04619499876 | *PCSK9* | Proprotein convertase subtilisin/kexin type 9 |
| OPGS001146 | 0.001611967462 | *SPATA20* | Spermatogenesis-associated protein 20 |
| OPGS001177 | -0.01157372997 | *GRP* | Gastrin-releasing peptide |
| OPGS001198 | -0.02194713069 | *RNASE1* | Ribonuclease pancreatic |
| OPGS001216 | -0.02385224655 | *MRC2* | C-type mannose receptor 2 |
| OPGS001223 | -0.02174759447 | *IL11RA* | Interleukin-11 receptor subunit alpha |
| OPGS001271 | 0.001163230837 | *LYVE1* | Lymphatic vessel endothelial hyaluronic acid receptor 1 |
| OPGS001297 | -0.003620619995 | *NEO1* | Neogenin |
| OPGS001313 | -0.07892920905 | *EREG* | Epiregulin |
| OPGS001342 | -0.02913239649 | *PCSK7* | Proprotein convertase subtilisin/kexin type 7 |
| OPGS001365 | -0.0871234396 | *CNMD* | Leukocyte cell-derived chemotaxin 1 |
| OPGS001375 | 0.02619559242 | *RARRES2* | Retinoic acid receptor responder protein 2 |
| OPGS001390 | -0.02485521964 | *RNASE2* | Non-secretory ribonuclease |
| OPGS001393 | 0.006674278744 | *LMAN2L* | VIP36-like protein |
| OPGS001402 | 0.01065824472 | *ARSB* | Arylsulfatase B |
| OPGS001415 | -0.02225537275 | *CCN6* | WNT1-inducible-signaling pathway protein 3 |
| OPGS001417 | 0.0217891806 | *SOD3* | Extracellular superoxide dismutase [Cu-Zn] |
| OPGS001433 | 0.01280237436 | *ARHGEF10* | Rho guanine nucleotide exchange factor 10 |
| OPGS001454 | 0.0005495793343 | *PTH2* | Tuberoinfundibular peptide of 39 residues |
| OPGS001501 | 0.01663983027 | *HBA1\|HBB* | Hemoglobin |
| OPGS001614 | 0.03479962306 | *CD14* | Monocyte differentiation antigen CD14 |
| OPGS001645 | 0.0349816372 | *GSTP1* | Glutathione S-transferase P |
| OPGS001665 | 0.05258538447 | *TREML1* | Trem-like transcript 1 protein |
| OPGS001701 | 0.02684681362 | *UAP1* | UDP-N-acetylhexosamine pyrophosphorylase |
| OPGS001713 | -0.002252387357 | *FBXO3* | F-box only protein 3 |
| OPGS001822 | -0.04812646106 | *LGALS9* | Galectin-9 |
| OPGS001867 | 0.03024654123 | *RELL1* | RELT-like protein 1 |
| OPGS001924 | -0.0753109244 | *LARGE1* | Glycosyltransferase-like protein LARGE1 |
| OPGS002035 | 0.01235344998 | *APBB1* | Amyloid beta A4 precursor protein-binding family B member 1 |
| OPGS002043 | 0.08974242284 | *MXRA8* | Matrix-remodeling-associated protein 8 |
| OPGS002083 | -0.04908492528 | *MSTN* | Growth/differentiation factor 8 |
| OPGS002092 | 0.03410393249 | *EXOSC1* | Exosome complex component CSL4 |
| OPGS002093 | -0.1117927878 | *FN1* | Fibronectin |
| OPGS002107 | 0.01880213061 | *SIGLEC1* | Sialoadhesin |
| OPGS002173 | 0.2016079898 | *IL2RB* | Interleukin-2 receptor subunit beta |

| | | | |
|---|---|---|---|
| OPGS002206 | 0.001430835757 | *GRID1* | Glutamate receptor ionotropic, delta-1 |
| OPGS002266 | 0.007828349338 | *CCS* | Copper chaperone for superoxide dismutase |
| OPGS002270 | -0.1996085367 | *TNFRSF21* | Tumor necrosis factor receptor superfamily member 21 |
| OPGS002297 | 0.003624469148 | *PRNP* | Major prion protein |
| OPGS002311 | -0.0599522431 | *OSCAR* | Osteoclast-associated immunoglobulin-like receptor |
| OPGS002327 | -5.93E-05 | *TAFA3* | Protein FAM19A3 |
| OPGS002342 | -0.05588214935 | *ERP29* | Endoplasmic reticulum resident protein 29 |
| OPGS002349 | 0.1319985638 | *SERPINA7* | Thyroxine-binding globulin |
| OPGS002375 | 0.08914472586 | *MMP2* | 72 kDa type IV collagenase |
| OPGS002387 | -0.03466151934 | *IL6R* | Interleukin-6 receptor subunit alpha |
| OPGS002422 | -0.009363639622 | *LTA* | Lymphotoxin-alpha |
| OPGS002432 | 0.00706934822 | *CTSS* | Cathepsin S |
| OPGS002435 | 0.02160488078 | *FGF5* | Fibroblast growth factor 5 |
| OPGS002452 | 0.0003270274597 | *MMP10* | Stromelysin-2 |
| OPGS002489 | -0.01226702235 | *SMOC2* | SPARC-related modular calcium-binding protein 2 |
| OPGS002493 | -0.01567557803 | *CXCL6* | C-X-C motif chemokine 6 |
| OPGS002496 | -0.01228166607 | *PON3* | Serum paraoxonase/lactonase 3 |
| OPGS002505 | 0.001052221987 | *TNFRSF10 B* | Tumor necrosis factor receptor superfamily member 10B |
| OPGS002538 | -0.009987184688 | *RGMA* | Repulsive guidance molecule A |
| OPGS002543 | 0.002872055009 | *UNC5C* | Netrin receptor UNC5C |
| OPGS002547 | 0.009615250715 | *THY1* | Thy-1 membrane glycoprotein |
| OPGS002558 | 0.01768499814 | *PCSK9* | Proprotein convertase subtilisin/kexin type 9 |
| OPGS002565 | 0.02950255496 | *IGFBP7* | Insulin-like growth factor-binding protein 7 |
| OPGS002566 | 0.02671122026 | *CEACAM8* | Carcinoembryonic antigen-related cell adhesion molecule 8 |
| OPGS002577 | 0.04296931942 | *IL1R1* | Interleukin-1 receptor type 1 |
| OPGS002581 | 0.02141441658 | *CX3CL1* | Fractalkine |
| OPGS002585 | 0.02036941018 | *CD5* | T-cell surface glycoprotein CD5 |
| OPGS002586 | -0.005057016904 | *DNER* | Delta and Notch-like epidermal growth factor-related receptor |
| OPGS002591 | 0.02039793342 | *SPP1* | Osteopontin |
| OPGS002602 | -0.01196488944 | *FABP2* | Fatty acid-binding protein, intestinal |
| OPGS002617 | 0.04411303598 | *CXCL9* | C-X-C motif chemokine 9 |
| OPGS002627 | 0.04394982516 | *CSF1* | Macrophage colony-stimulating factor 1 |
| OPGS002640 | -0.08018002738 | *NTRK3* | NT-3 growth factor receptor |
| OPGS002645 | 0.0122790329 | *CPA1* | Carboxypeptidase A1 |
| OPGS002646 | -0.05376216225 | *HBEGF* | Proheparin-binding EGF-like growth factor |
| OPGS002663 | 0.2108586171 | *CLEC1B* | C-type lectin domain family 1 member B |
| OPGS002689 | -0.01341380662 | *ITGB1BP2* | Integrin beta-1-binding protein 2 |

**Supplementary Table 16** Genetic risk score for 142 metabolites were retained in the MetPRS lasso models for CAD prediction

| OMICSPRED_ID | Weight | Metabolite |
|---|---|---|
| OPGS002724 | 0.01064543107 | imidazole lactate |
| OPGS002726 | -0.0007380892243 | methionylalanine |
| OPGS002732 | 0.02228577071 | escitalopram |
| OPGS002735 | 0.002193451725 | etiocholanolone glucuronide |
| OPGS002741 | 0.0005853793621 | methionine sulfone |
| OPGS002753 | 0.008032904951 | andro steroid monosulfate (1)* |
| OPGS002763 | -0.0236687058 | dimethylglycine |
| OPGS002767 | -0.003884915037 | phenylalanylarginine |
| OPGS002772 | -0.01506523739 | N-acetylcarnosine |
| OPGS002776 | 0.01337189577 | aspartate |
| OPGS002779 | 0.002963817688 | X - 21470 |
| OPGS002785 | 0.00667089087 | flavin adenine dinucleotide (FAD) |
| OPGS002797 | -0.03723528546 | 5-hydroxylysine |
| OPGS002800 | 0.006262642555 | methyl glucopyranoside (alpha + beta) |
| OPGS002814 | -0.003494537117 | spermidine |
| OPGS002818 | 0.01495707245 | X - 24241 |
| OPGS002824 | 0.007295977116 | 4-acetamidobutanoate |
| OPGS002837 | 0.001245801916 | 1-stearoyl-2-arachidonoyl-GPI (18:0/20:4) |
| OPGS002840 | 0.02592871189 | X - 18914 |
| OPGS002841 | -0.04889035702 | sphingomyelin (d18:1/20:1, d18:2/20:0)* |
| OPGS002842 | 0.01218441792 | glycosyl-N-palmitoyl-sphingosine |
| OPGS002849 | -0.03359799822 | X - 16071 |
| OPGS002853 | 0.02639529214 | orotate |
| OPGS002854 | -0.01351156176 | oleoyl ethanolamide |
| OPGS002865 | -5.95E-05 | X - 21283 |
| OPGS002866 | 0.01969024358 | N-acetyltryptophan |
| OPGS002871 | 0.02197677148 | X - 12459 |
| OPGS002874 | 0.0119875922 | campesterol |
| OPGS002876 | 0.003236446726 | 3-methoxytyrosine |
| OPGS002877 | -0.004389426674 | 2-hydroxyibuprofen |
| OPGS002886 | 0.02133896815 | X - 12822 |
| OPGS002887 | -0.0003973123569 | X - 11308 |
| OPGS002889 | 0.008758564135 | N6-acetyllysine |
| OPGS002897 | 0.0001289778044 | 6-oxopiperidine-2-carboxylic acid |
| OPGS002901 | -0.05211785059 | X - 23369 |
| OPGS002902 | 0.01842405613 | X - 23749 |
| OPGS002904 | 0.03221575502 | 1-arachidonoyl-GPI (20:4)* |
| OPGS002909 | -0.001558981717 | X - 24439 |
| OPGS002910 | -0.005136801805 | X - 24422 |
| OPGS002911 | -0.006648872452 | 1-methylurate |
| OPGS002914 | -0.0267239753 | N-acetyl-beta-alanine |
| OPGS002919 | 0.00144888733 | cysteine-glutathione disulfide |
| OPGS002939 | 0.01221925475 | cis-4-decenoyl carnitine |
| OPGS002940 | 0.005245773472 | lactosyl-N-palmitoyl-sphingosine |
| OPGS002950 | 0.002838844091 | beta-hydroxyisovalerate |
| OPGS002956 | 0.0377711717 | sphingomyelin (d18:1/15:0, d16:1/17:0)* |

| | | |
|---|---|---|
| OPGS002974 | 0.005277437713 | N6,N6,N6-trimethyllysine |
| OPGS002988 | -0.02258601577 | 2-stearoyl-GPE (18:0)* |
| OPGS002990 | 0.02325484405 | 10-undecenoate (11:1n1) |
| OPGS002994 | 0.05029787654 | glycohyocholate |
| OPGS002995 | -0.1236514015 | 5alpha-androstan-3alpha,17beta-diol disulfate |
| OPGS002996 | 0.1210693333 | cystathionine |
| OPGS003007 | -0.001860366292 | trans-urocanate |
| OPGS003024 | 0.003341430858 | X - 13529 |
| OPGS003030 | 0.08149119016 | 1-methylhistidine |
| OPGS003032 | -0.02248923193 | N-acetylalanine |
| OPGS003038 | -0.03437870141 | 1-margaroyl-2-linoleoyl-GPC (17:0/18:2)* |
| OPGS003041 | 0.1953635966 | 1-oleoyl-2-linoleoyl-glycerol (18:1/18:2) |
| OPGS003053 | 0.03326932939 | taurolithocholate 3-sulfate |
| OPGS003058 | 0.00282201535 | sphingomyelin (d18:1/17:0, d17:1/18:0, d19:1/16:0) |
| OPGS003065 | 0.01736624591 | X - 12100 |
| OPGS003066 | 0.03399558413 | ribonate |
| OPGS003080 | 0.005985689146 | S-methylcysteine |
| OPGS003081 | 0.00236466083 | DSGEGDFXAEGGGVR* |
| OPGS003097 | -0.05471657953 | X - 12216 |
| OPGS003104 | -0.07240839096 | indoleacetate |
| OPGS003112 | 0.02984216205 | sarcosine (N-Methylglycine) |
| OPGS003115 | 0.05360666324 | sphingosine 1-phosphate |
| OPGS003119 | 0.07615086419 | 1-palmitoyl-GPC (16:0) |
| OPGS003140 | -0.002977670106 | ethyl glucuronide |
| OPGS003143 | -0.002945373901 | 3-(4-hydroxyphenyl)lactate |
| OPGS003144 | 0.01598018546 | X - 12026 |
| OPGS003145 | 0.03242404389 | adenosine 3',5'-cyclic monophosphate (cAMP) |
| OPGS003149 | 0.05850394618 | sphingosine |
| OPGS003158 | -0.09108521262 | 2-hydroxy-3-methylvalerate |
| OPGS003166 | 0.0001216524 | thyroxine |
| OPGS003176 | 0.01111436297 | X - 12442 |
| OPGS003179 | 0.0252562106 | 4-methyl-2-oxopentanoate |
| OPGS003183 | -0.08010570018 | X - 16944 |
| OPGS003186 | -0.05630484334 | arabitol/xylitol |
| OPGS003195 | 0.1239971815 | indole-3-carboxylic acid |
| OPGS003197 | 0.1299638644 | 1-stearoyl-GPC (18:0) |
| OPGS003200 | -0.02344664131 | threonine |
| OPGS003204 | 0.01147518027 | X - 18886 |
| OPGS003219 | -0.04062021573 | N-acetylvaline |
| OPGS003244 | 0.02116186576 | 3-hydroxybutyrylcarnitine (1) |
| OPGS003250 | 0.05370210276 | X - 22850 |
| OPGS003263 | 0.03398495105 | 1,3,7-trimethylurate |
| OPGS003275 | -0.02370492344 | 2-linoleoyl-GPC (18:2)* |
| OPGS003278 | -0.05480669248 | pregnanolone/allopregnanolone sulfate |
| OPGS003283 | -0.1253347361 | N-acetylthreonine |
| OPGS003287 | -0.01892922506 | isoeugenol sulfate |
| OPGS003289 | -0.05951829922 | X - 12170 |
| OPGS003296 | -0.0596992726 | X - 24456 |

| | | |
|---|---|---|
| OPGS003300 | -0.03082724168 | choline |
| OPGS003301 | 0.1644810923 | pseudouridine |
| OPGS003303 | 0.06935890086 | X - 24240 |
| OPGS003305 | -0.04508444024 | 7-alpha-hydroxy-3-oxo-4-cholestenoate (7-Hoca) |
| OPGS003306 | 0.02458500613 | 1-stearoyl-2-oleoyl-GPC (18:0/18:1) |
| OPGS003326 | -0.03581523741 | galactonate |
| OPGS003352 | -0.0844024952 | piperine |
| OPGS003353 | -0.04388267615 | X - 16580 |
| OPGS003356 | 0.05233836142 | 1-linoleoylglycerol (18:2) |
| OPGS003358 | 0.1187753095 | pyridoxate |
| OPGS003360 | 0.08173477267 | eugenol sulfate |
| OPGS003369 | -0.05314215624 | 9,10-DiHOME |
| OPGS003372 | 0.1007701944 | X - 12206 |
| OPGS003375 | 0.2316478529 | p-cresol sulfate |
| OPGS003377 | -0.008380219927 | caprate (10:0) |
| OPGS003383 | 0.01372398979 | 3-methylglutaconate |
| OPGS003386 | -0.02735246311 | pyroglutamylvaline |
| OPGS003391 | -0.1642487936 | palmitoleate (16:1n7) |
| OPGS003392 | -0.1449473242 | tiglylcarnitine |
| OPGS003394 | 0.03125613459 | 3-hydroxybutyrate (BHBA) |
| OPGS003396 | -0.1407631469 | erucate (22:1n9) |
| OPGS003397 | -0.05619027155 | X - 21666 |
| OPGS003401 | -0.07126300271 | 1-stearoyl-GPI (18:0) |
| OPGS003405 | 0.08029825439 | taurocholate |
| OPGS003406 | 0.003903562919 | sphingomyelin (d18:2/24:1, d18:1/24:2)* |
| OPGS003411 | 0.0717473073 | X - 24307 |
| OPGS003420 | 0.00480877013 | Cholesterol in small LDL |
| OPGS003425 | 0.00556437064 | Cholesterol in large LDL |
| OPGS003429 | 0.03155959373 | Cholesteryl esters in large LDL |
| OPGS003430 | 0.02737687011 | Concentration of small LDL particles |
| OPGS003436 | 0.1127892428 | Phospholipids in large LDL |
| OPGS003444 | 0.03169083868 | Apolipoprotein B |
| OPGS003445 | -0.01962962426 | Glycine |
| OPGS003447 | -0.02935890684 | Average diameter for HDL particles |
| OPGS003450 | 0.009270879439 | Triglycerides in IDL |
| OPGS003453 | 0.001502308033 | Triglycerides in very small VLDL |
| OPGS003459 | -0.09216171595 | Cholesterol in large HDL |
| OPGS003463 | 0.1375086676 | Phospholipids in small VLDL |
| OPGS003470 | 0.08431859857 | Triglycerides in medium LDL |
| OPGS003482 | -0.1790821557 | Triglycerides in chylomicrons and extremely large VLDL |
| OPGS003489 | -0.128728054 | Concentration of chylomicrons and extremely large VLDL particles |
| OPGS003508 | 0.04901813304 | Triglycerides in small HDL |
| OPGS003510 | -0.06609015174 | Cholesterol in chylomicrons and extremely large VLDL |
| OPGS003527 | -0.06658381994 | Sphingomyelins |
| OPGS003531 | 0.01159311555 | Cholesteryl esters in small HDL |
| OPGS003555 | -0.0938082915 | Isoleucine |
| OPGS003557 | 0.4937141706 | Glucose |

| | | |
|---|---|---|
| OPGS003558 | -0.004902122593 | Acetoacetate |

**Supplementary Table 17** Familial hypercholesterolemia variants and their effects on low-density lipoprotein cholesterol

| Variant | Gene | #Carriers | Beta (LDL-c) | Consequence | Amino acid or cDNA change |
|---|---|---|---|---|---|
| 1:55057454:G>T | PCSK9 | 2 | 2.968 | missense_variant | p.Asp374Tyr |
| 1:55058630:C>T | PCSK9 | 41 | 1.153 | missense_variant | p.Arg496Trp |
| 2:21006288:C>T | APOB | 343 | 1.457 | missense_variant | p.Arg3527Gln |
| 2:21006289:G>A | APOB | 16 | 0.632 | missense_variant | p.Arg3527Trp |
| 19:11100272:CA>C | LDLR | 2 | 1.071 | frameshift_variant | g.11100273del |
| 19:11100291:T>G | LDLR | 6 | 1.755 | missense_variant | p.Cys131Gly |
| 19:11102714:C>T | LDLR | 6 | 1.940 | missense_variant | p.Arg166Cys |
| 19:11102732:T>G | LDLR | 16 | 1.611 | missense_variant | p.Trp172Gly |
| 19:11102739:G>A | LDLR | 6 | 1.938 | missense_variant | p.Cys174Tyr |
| 19:11102774:G>A | LDLR | 34 | 2.576 | missense_variant | p.Glu186Lys |
| 19:11102787:G>A | LDLR | 23 | 2.296 | splice_donor_variant | g.11102787G>A |
| 19:11102787:G>C | LDLR | 2 | 0.743 | splice_donor_variant | g.11102787G>C |
| 19:11102787:G>GT | LDLR | 2 | 0.364 | splice_donor_variant | g.11102788ins |
| 19:11105243:G>A | LDLR | 39 | 0.946 | missense_variant | p.Glu198Lys |
| 19:11105249:C>T | LDLR | 4 | 0.875 | missense_variant | p.Arg200Cys |
| 19:11105268:G>T | LDLR | 2 | 1.561 | missense_variant | p.Cys206Phe |
| 19:11105324:G>A | LDLR | 2 | 1.164 | missense_variant | p.Glu225Lys |
| 19:11105337:C>T | LDLR | 4 | 0.996 | missense_variant | p.Pro229Leu |
| 19:11105407:C>A | LDLR | 2 | 1.497 | stop_gained | p.Cys252* |
| 19:11105408:G>A | LDLR | 45 | 1.083 | missense_variant | p.Asp253Asn |
| 19:11105415:AC>A | LDLR | 3 | 2.850 | frameshift_variant | g.11105416del |
| 19:11105448:C>T | LDLR | 6 | 0.553 | missense_variant | p.Pro266Leu |
| 19:11105470:C>G | LDLR | 3 | 2.655 | stop_gained | p.Tyr273* |
| 19:11105567:G>A | LDLR | 3 | 2.298 | missense_variant | p.Asp306Asn |
| 19:11105568:A>G | LDLR | 15 | 1.363 | missense_variant | p.Asp306Gly |
| 19:11105585:GAC>G | LDLR | 6 | 1.926 | frameshift_variant | g.11105586_11105587del |
| 19:11105587:C>G | LDLR | 10 | 2.036 | missense_variant | p.Asp312Glu |
| 19:11105588:G>C | LDLR | 2 | 3.688 | missense_variant | p.Glu313Gln |
| 19:11105588:G>T | LDLR | 5 | 1.040 | stop_gained | p.Glu313* |
| 19:11106588:G>A | LDLR | 50 | 2.759 | missense_variant | p.Glu325Lys |
| 19:11106631:A>C | LDLR | 6 | 1.136 | missense_variant | p.Gln339Pro |
| 19:11107436:G>A | LDLR | 24 | 0.393 | missense_variant | p.Glu373Lys |
| 19:11107461:G>A | LDLR | 1 | 3.669 | missense_variant | p.Cys381Tyr |
| 19:11107486:C>G | LDLR | 8 | 1.125 | missense_variant | p.Asp389Glu |
| 19:11107506:A>C | LDLR | 55 | 0.794 | missense_variant | p.Lys396Thr |
| 19:11107512:G>A | LDLR | 12 | 2.058 | missense_variant | p.Cys398Tyr |
| 19:11110714:G>A | LDLR | 9 | 0.387 | missense_variant | p.Gly420Ser |
| 19:11110759:C>T | LDLR | 8 | 2.773 | stop_gained | p.Arg435* |
| 19:11111571:G>A | LDLR | 2 | 1.702 | missense_variant | p.Gly458Asp |
| 19:11113268:G>A | LDLR | 4 | 0.596 | splice_polypyrimidine_tract_variant | g.11113268G>A |
| 19:11113287:C>A | LDLR | 4 | 0.783 | missense_variant | p.Ala484Val |
| 19:11113295:TTC>T | LDLR | 4 | 1.751 | frameshift_variant | g.11113297_11113298del |
| 19:11113307:C>T | LDLR | 12 | 1.248 | missense_variant | p.Arg491Trp |
| 19:11113308:G>A | LDLR | 6 | 0.745 | missense_variant | p.Arg491Gln |
| 19:11113313:G>A | LDLR | 2 | 1.269 | missense_variant | p.Glu493Lys |
| 19:11113322:A>G | LDLR | 1 | 2.247 | missense_variant | p.Lys496Glu |
| 19:11113329:C>T | LDLR | 51 | 0.890 | missense_variant | p.Thr498Met |
| 19:11113337:C>T | LDLR | 7 | 1.805 | missense_variant | p.Arg501Trp |
| 19:11113554:CA>C | LDLR | 1 | 0.748 | frameshift_variant | g.11113555del |
| 19:11113590:G>T | LDLR | 15 | 0.714 | missense_variant | p.Asp557Tyr |
| 19:11113612:T>C | LDLR | 11 | 2.290 | missense_variant | p.Leu564Pro |
| 19:11113620:G>A | LDLR | 105 | 1.461 | missense_variant | p.Asp567Asn |

| | | | | | |
|---|---|---|---|---|---|
| 19:11113650:G>A | LDLR | 1 | 0.054 | missense_variant | p.Asp577Asn |
| 19:11113678:C>T | LDLR | 8 | 1.189 | missense_variant | p.Ala586Val |
| 19:11113743:G>A | LDLR | 5 | 1.891 | missense_variant | p.Val608Met |
| 19:11116114:G>A | LDLR | 2 | 3.731 | stop_gained | p.Trp621* |
| 19:11116125:G>A | LDLR | 4 | 1.346 | missense_variant | p.Ala625Thr |
| 19:11116139:AG>A | LDLR | 2 | 1.637 | frameshift_variant | g.11116140del |
| 19:11116141:G>A | LDLR | 8 | 1.695 | missense_variant | p.Gly630Glu |
| 19:11116198:A>G | LDLR | 4 | 0.621 | missense_variant | p.Asn649Ser |
| 19:11116873:C>T | LDLR | 10 | 0.544 | missense_variant | p.Arg659Cys |
| 19:11116898:T>C | LDLR | 4 | 1.902 | missense_variant | p.Leu667Pro |
| 19:11116928:G>A | LDLR | 2 | 2.506 | missense_variant | p.Gly677Glu |
| 19:11116936:C>T | LDLR | 16 | 1.425 | missense_variant | p.Arg680Trp |
| 19:11116970:C>A | LDLR | 42 | 0.590 | missense_variant | p.Ala691Asp |
| 19:11116976:C>G | LDLR | 1 | 5.343 | missense_variant | p.Pro693Arg |
| 19:11120091:G>A | LDLR | 3 | 0.861 | splice_acceptor_variant | g.11120091G>A |
| 19:11120110:GAT>G | LDLR | 1 | 2.137 | frameshift_variant | g.11120111_11120112del |
| 19:11120143:C>T | LDLR | 37 | 0.809 | missense_variant | p.Arg718Cys |
| 19:11120144:G>A | LDLR | 5 | 0.913 | missense_variant | p.Arg718His |
| 19:11120152:G>A | LDLR | 10 | 0.416 | missense_variant | p.Gly721Cys |
| 19:11120370:G>A | LDLR | 1 | 0.391 | missense_variant | p.Gly748Glu |
| 19:11120436:C>T | LDLR | 37 | 2.101 | missense_variant | p.Pro770Leu |
| 19:11120442:T>TC | LDLR | 3 | 4.052 | frameshift_variant | g.11120443ins |
| 19:11120501:G>A | LDLR | 75 | 0.139 | missense_variant | p.Asp792Asn |
| 19:11123200:G>T | LDLR | 3 | 1.366 | stop_gained | p.Glu808* |
| 19:11123324:TA>T | LDLR | 1 | 2.237 | frameshift_variant | g.11123325del |
| 19:11128027:C>CA | LDLR | 1 | 1.884 | frameshift_variant | g.11128028ins |

Genome positions use GRCh38 coordinates. The number of carriers was directly counted from the UK Biobank whole exome sequence (WES) data and the betas for LDL-c were estimated from an iterative conditional regression analysis by the software Regenie v3.3.

**Supplementary Table 18** Definition of coronary artery disease in UK Biobank

| DataField | DataFiled explained | Coding |
|---|---|---|
| 20002 | Non-cancer illness code, self-reported | 1075 |
| 20004 | Operation code, self-reported | 1070,1095,1523 |
| 41203 | Diagnoses - main ICD9 | 410,4109,411,4119,412,4129,4140,4148,4149 |
| 41205 | Diagnoses - secondary ICD9 | 410,4109,411,4119,412,4129,4140,4148,4149 |
| 41271 | Diagnoses - ICD9 | 410,4109,411,4119,412,4129,4140,4148,4149 |
| 41202 | Diagnoses - main ICD10 | I21,I21.0,I21.1,I21.2,I21.3,I21.4,I21.9,I22,I22.0,I22.1,I22.8,I22.9,I23,I23.0,I23.1,I23.2,I23.3,I23.4,I23.5,I23.6,I23.8,I24.0,I24.1,I24.8,I24.9,I25.0,I25.1,I25.2,I25.5,I25.6,I25.8,I25.9 |
| 41204 | Diagnoses - secondary ICD10 | I21,I21.0,I21.1,I21.2,I21.3,I21.4,I21.9,I22,I22.0,I22.1,I22.8,I22.9,I23,I23.0,I23.1,I23.2,I23.3,I23.4,I23.5,I23.6,I23.8,I24.0,I24.1,I24.8,I24.9,I25.0,I25.1,I25.2,I25.5,I25.6,I25.8,I25.9 |
| 41270 | Diagnoses - ICD10 | I21,I21.0,I21.1,I21.2,I21.3,I21.4,I21.9,I22,I22.0,I22.1,I22.8,I22.9,I23,I23.0,I23.1,I23.2,I23.3,I23.4,I23.5,I23.6,I23.8,I24.0,I24.1,I24.8,I24.9,I25.0,I25.1,I25.2,I25.5,I25.6,I25.8,I25.9 |
| 40001 | Underlying (primary) cause of death: ICD10 | I21,I21.0,I21.1,I21.2,I21.3,I21.4,I21.9,I22,I22.0,I22.1,I22.8,I22.9,I23,I23.0,I23.1,I23.2,I23.3,I23.4,I23.5,I23.6,I23.8,I24.0,I24.1,I24.8,I24.9,I25.0,I25.1,I25.2,I25.5,I25.6,I25.8,I25.9 |
| 40002 | Contributory (secondary) causes of death: ICD10 | I21,I21.0,I21.1,I21.2,I21.3,I21.4,I21.9,I22,I22.0,I22.1,I22.8,I22.9,I23,I23.0,I23.1,I23.2,I23.3,I23.4,I23.5,I23.6,I23.8,I24.0,I24.1,I24.8,I24.9,I25.0,I25.1,I25.2,I25.5,I25.6,I25.8,I25.9 |
| 41200 | Operative procedures - main OPCS4 | K49,K49.1,K49.2,K49.3,K49.4,K49.8,K49.9,K75,K75.1,K75.2,K75.3,K75.4,K75.8,K75.9,K50.1,K50.2,K50.4,K40,K40.1,K40.2,K40.3,K40.4,K40.8,K40.9,K41,K41.1,K41.2,K41.3,K41.4,K41.8,K41.9,K42,K42.1,K42.2,K42.3,K42.4,K42.8,K42.9,K43,K43.1,K43.2,K43.3,K43.4,K43.8,K43.9,K44,K44.1,K44.2,K44.8,K44.9,K45.1,K45.2,K45.3,K45.4,K45.5,K45.6,K45.8,K45.9,K46,K46.1,K46.2,K46.3,K46.4,K46.5,K46.8,K46.9,K47.1 |
| 41210 | Operative procedures - secondary OPCS4 | K49,K49.1,K49.2,K49.3,K49.4,K49.8,K49.9,K75,K75.1,K75.2,K75.3,K75.4,K75.8,K75.9,K50.1,K50.2,K50.4,K40,K40.1,K40.2,K40.3,K40.4,K40.8,K40.9,K41,K41.1,K41.2,K41.3,K41.4,K41.8,K41.9,K42,K42.1,K42.2,K42.3,K42.4,K42.8,K42.9,K43,K43.1,K43.2,K43.3,K43.4,K43.8,K43.9,K44,K44.1,K44.2,K44.8,K44.9,K45.1,K45.2,K45.3,K45.4,K45.5,K45.6,K45.8,K45.9,K46,K46.1,K46.2,K46.3,K46.4,K46.5,K46.8,K46.9,K47.1 |

**Supplementary Table 19** Lipid values adjustment rationale

| Medication | Lipid Values Adjustment | | | | Reference(s) |
|---|---|---|---|---|---|
| | Total Cholesterol | LDL | HDL | Triglycerides | |
| **Statin** | -20% | -30% | NA | -15% | Cholesterol Treatment Trialists' (CTT) Collaboration et al. Lancet. 2010 Nov 13;376(9753) |
| **Ezetimibe** | -15% | -20% | NA | NA | 1- Sudhop T et al. Circulation 2002;106:1943-1948<br>2- Cannon CP et al. NEJM 2015;372:2387-97<br>3- Zhao Z et al. Medicine (Baltimore). 2019 Feb;98(6) |
| **Bile Acid Sequestrant** | NA | -15% | NA | NA | Lloyd-Jones et al. JACC 2017;70(14) |
| **Fibrate** | -10% | -10% | +10% | -35% | Birjmohun RS et al. JACC 2005;45:185-97 |
| **Niacin** | -10% | -10% | +15% | -20% | Birjmohun RS et al. JACC 2005;45:185-97 |
| **Not specified** | -20% | -30% | NA | -15% | Assumed statin |

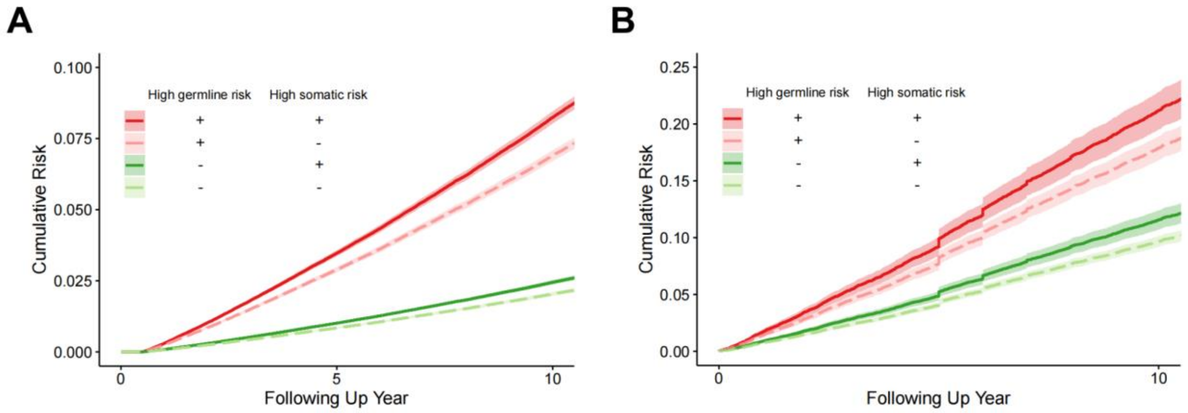


**Supplementary Fig. 1**. Cumulative risks of CAD for combinations of GermRisk and SomaRisk in (A) UK Biobank and (B) TOPMed.

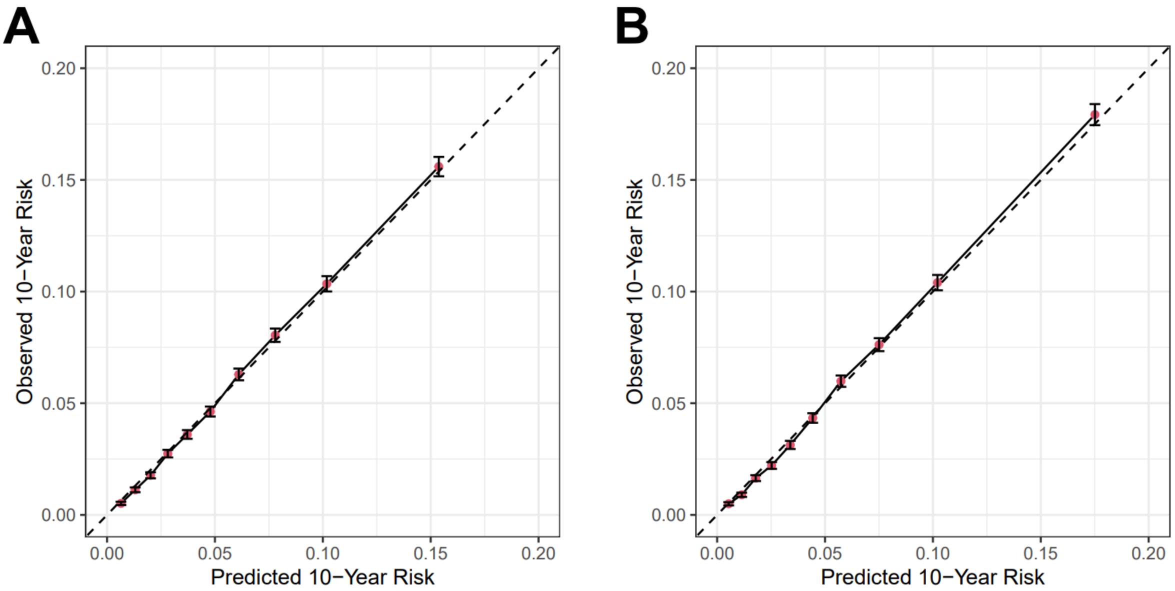


**Supplementary Fig. 2**. Calibration plot for (A) PCE and (B) PCE + IGM. Calibration was performed by R package rms(v6.7-1) with parameters time point u=10, group predicted containing samples m=37728, number of bootstrapping to estimate the confidence interval for observed risk B=100, and the method for validating survival predictions method='KM' (Kaplan-Meier estimates). IGM: integrated genomic model; PCE, pooled cohort equation.

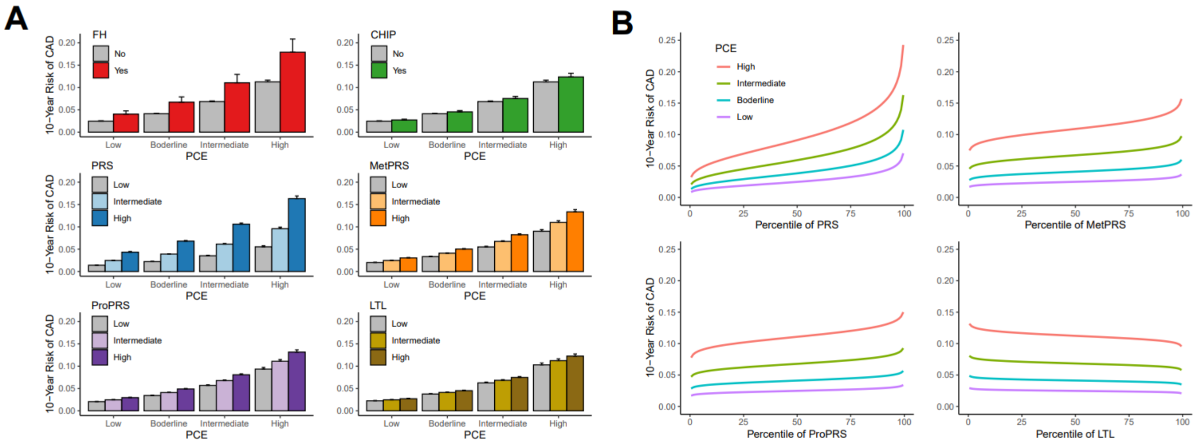


**Supplementary Fig. 3**. (A) Risk stratification by genetic drivers within each PCE risk stratum. (B) Predicted 10-year CAD risk gradient by genetic risk percentile. IGM categories were defined as low risk (bottom 20%), intermediate risk (middle 60%), and high risk group (top 20%), respectively. PCE categories were defined as low (estimated risk less than 5%), borderline (risk between 5% to 7.5%), intermediate (risk between 7.5% and 20%), and high (risk greater than 20%), respectively. IGM: integrated genomic model; PCE, pooled cohort equation.

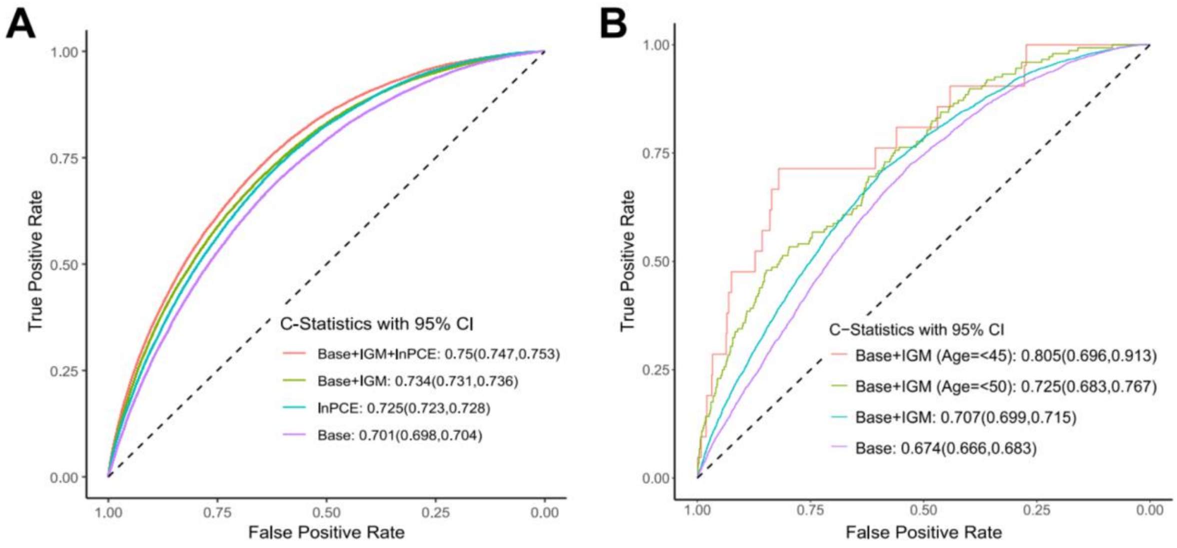


**Supplementary Fig. 4.** Receiver operator characteristic curves and C-statistic for different models in (A) UK Biobank and (B) TOPMed. Base model: age, sex, top 10 principal components. IGM (integrated genomic model) with six drivers including FH, PRS, MetPRS, ProPRS, CHIP, and LTL. PCE, pooled cohort equation. lnPCE: log transformed PCE.

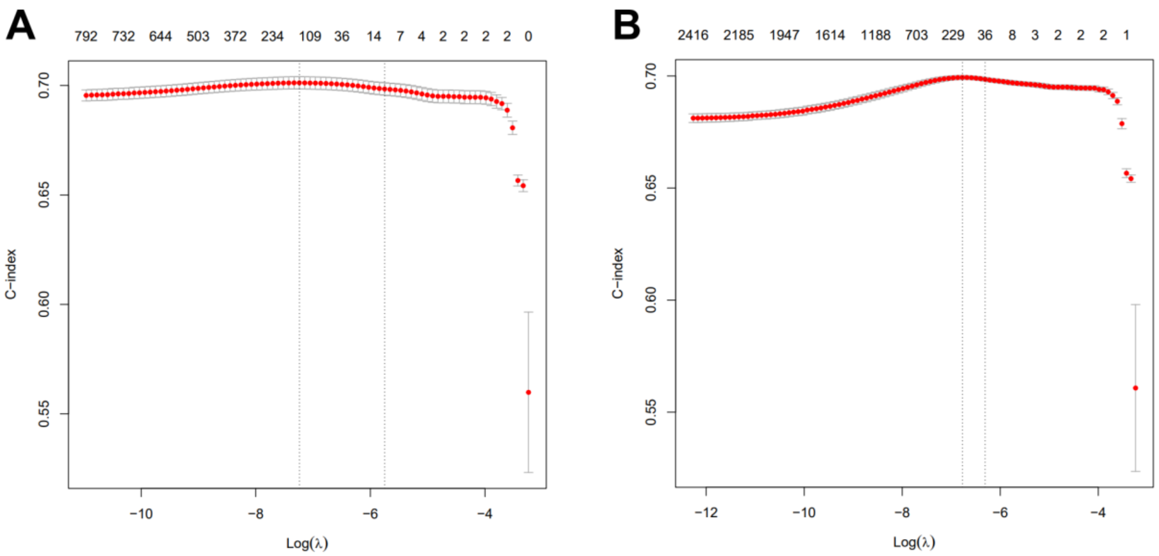


**Supplementary Fig. 5.** Five-fold cross validation to choose the best lambda in the training process (A) MetPRS and (B) ProPRS via Lasso, using Harrell's concordance (C-index) as performance measurement.

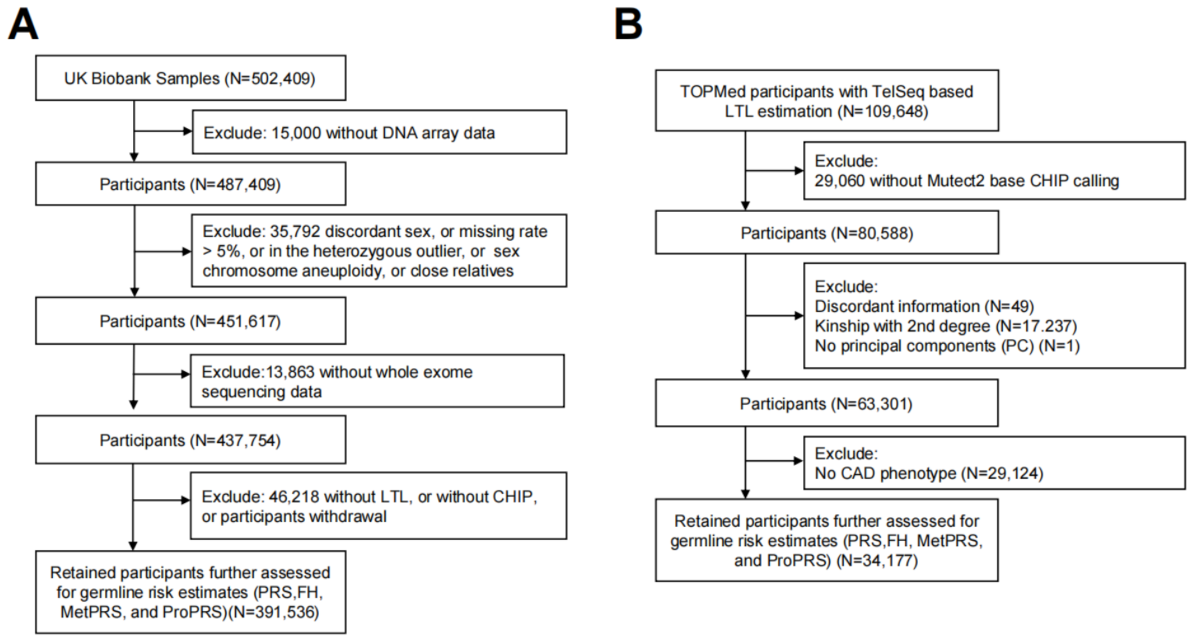


**Supplementary Fig. 6.** Quality control procedures of study participants in (A) UK Biobank and (B) TOPMed.

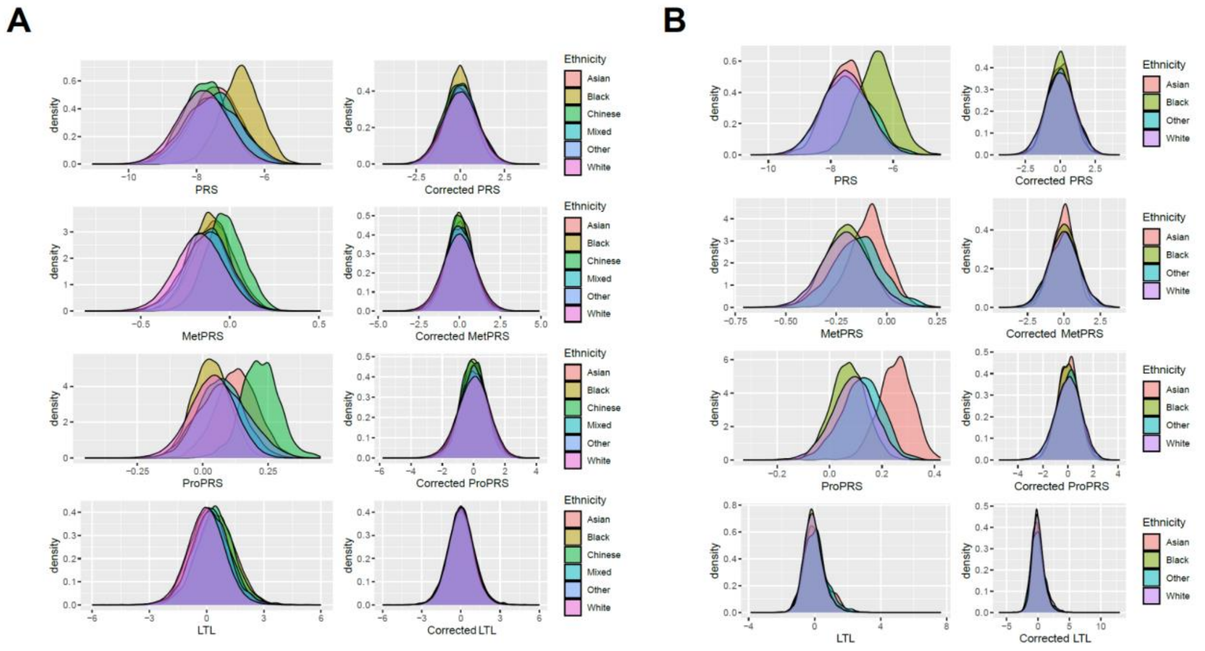


**Supplementary Fig. 7.** Distribution of genetic drivers across ancestries before and after adjusting for age, sex, and top 10 PCs in (A) UK Biobank and (B) TOPMed.
